# Mitochondrial activity promotes neutrophil degranulation and endothelial dysfunction in systemic infections

**DOI:** 10.1101/2025.10.10.25337737

**Authors:** Przemysław Zakrzewski, Christopher M. Rice, Claire Naveh, Isaac Dowell, Kathryn Fleming, Aravind V. Ramesh, Rachel Jones, Pedro L. Moura, Drinalda Cela, Sarah Groves, Stephanie Fletcher-Jones, Yohance Victory, Mainga Bhima, Stefan Ebmeier, Laura Carey, Matthew Butler, Simon C. Satchell, Ase Berg, Nadia Palolite, James Nyirenda, Watipenge Nyasulu, Isabel Zgambo, Charalampos Attipa, Linda Wooldridge, Andrew Davidson, Aubrey Cunnington, Christopher A. Moxon, Borko Amulic

**Author notes:** Equal contribution (Przemysław Zakrzewski, Christopher M. Rice, Claire Naveh). Equal contribution (Christopher A. Moxon, Borko Amulic).

## Abstract

Neutrophils are essential for defense against pathogens but excessive activation in systemic infections can drive immunopathology. We show that neutrophil degranulation can induce endothelial dysfunction via degradation of the glycocalyx and increase of endothelial permeability. To identify targetable pathways regulating neutrophil degranulation in severe inflammation, we compared the proteomes of neutrophils isolated from patients with severe malaria and sepsis. We found significant upregulation of mitochondrial pathways, which was accompanied by increased rates of mitochondrial respiration and was linked to neutrophil immaturity. Malaria induced mitochondrial fusion and networking, while sepsis was associated with mitochondrial biogenesis. Immature neutrophils in both infections produced elevated levels of mitochondrial ROS, which enhanced release of primary and secondary granules. Our study provides a mechanistic explanation for the hyperinflammatory nature of immature neutrophils and points to pharmacological scavenging of mitochondrial ROS as a potential therapeutic strategy to reduce endothelial damage in severe inflammation.

## INTRODUCTION

Regulation of neutrophil abundance and activity is essential for optimal health. Produced at a rate of up to 100 billion per day [1], neutrophils patrol the vasculature and suppress pathogens by phagocytosis, degranulation and release of antimicrobial chromatin in the form of neutrophil extracellular traps (NETs) [2]. Yet the role of neutrophils in infection is complex: efficient microbial clearance needs to be carefully balanced with containment of host tissue damage. This is particularly prominent in systemic infections such as severe malaria and sepsis, where neutrophil activation can have cytotoxic effects on endothelial cells, driving vascular breakdown and promoting multi-organ failure [3–5]. Targeting neutrophils is a promising strategy to reverse endothelial dysfunction, a therapeutic approach that is of considerable interest in multiple vascular diseases.

In *Plasmodium falciparum* (*P. falciparum*) malaria, infected erythrocytes cytoadhere to microvascular endothelial beds via parasite-encoded cytoadhesion factors [6], which bind endothelial receptors embedded in the vascular glycocalyx [4, 7], a carbohydrate rich layer coating the luminal side of endothelial cells. This initiates endothelial activation, a process that is enhanced by inflammatory factors, leading to vascular leak and oedema [7]. For reasons that are poorly understood, in some individuals a combination of systemic inflammation and tissue-specific parasite adhesion can result in damage to organs such as brain and lung [6], leading to coma and respiratory distress. Neutrophils are thought to have direct and indirect anti-parasitic activity [3, 8–10]. On the other hand, multiple transcriptomic studies identified a neutrophil signature in patient blood that discriminates between uncomplicated and severe malaria [11–13]. Cerebral malaria is associated with elevated plasma concentrations of neutrophil granule proteins [14] [15], while NETs colocalize with damaged endothelium in cerebral malaria autopsy studies [16, 17]. Neutrophil serine proteases have been shown to promote immunopathology in mouse malaria models by enhancing parasite sequestration and via direct cytotoxic effects [17, 18].

In sepsis, neutrophils similarly have dual roles in microbial clearance and in promoting endothelial injury and multi-organ failure [5]. Although the pathophysiology of sepsis is complex, in the hyperinflammatory stage, neutrophils can damage the glycocalyx [19, 20] and induce barrier breakdown [21]. This promotes vascular leak and enhances inflammatory cell recruitment and extravasation, which are all steps in progression to multiorgan dysfunction syndrome [22, 23]. As in malaria, neutrophil proteases are implicated in endothelial dysfunction and poor outcome in sepsis [24, 25].

In order to target neutrophils in vascular inflammation, we need a better understanding of neutrophil activation and whether all neutrophil states have equal propensity to cause endothelial damage. In particular, it remains unclear whether systemic inflammation leads to qualitative changes in neutrophil development and how this impacts effector responses. Neutrophil maturation state may be a factor in determining inflammatory potential; the emergence of immature circulating neutrophils has been reported in many inflammatory diseases, including malaria [26], sepsis [27], severe COVID-19 [28] and systemic lupus erythematosus [29]. These studies suggest that immature neutrophils have altered or heightened effector functions, which can damage host tissues [30, 31]. The molecular or developmental mechanisms underpinning such altered responses remain unknown, although it has been suggested that altered metabolism plays a role [32, 33].

We hypothesized that developmental changes in granulopoiesis caused by severe inflammation may promote immunopathology. Comparing the proteomes of neutrophils isolated from patients with severe malaria and sepsis, we identified mitochondrial metabolism and mitochondrial reactive oxygen species (ROS) production as shared pathways regulating exocytosis of cytotoxic neutrophil proteins.

## RESULTS

### Primary granule proteins enhance endothelial permeability and glycocalyx shedding

To confirm the potential of human neutrophils to damage the endothelium, we investigated their ability to degrade the glycocalyx, a vasculoprotective layer of glycosaminoglycans and proteoglycans that maintains an anti-inflammatory state and preserves endothelial barrier integrity [34]. As a model, we used glomerular endothelial cells (GEnCs), a kidney microvascular endothelial cell line [35] with an abundant glycocalyx. Degradation of the glycocalyx on cultured endothelial cells causes release of the syndecan-4 proteoglycan into the supernatant, which can be quantified by ELISA as a readout of glycocalyx damage. Co-incubation of GEnCs with naive neutrophils induced only minimal glycocalyx shedding (Fig. 1A); however, co-incubation with neutrophils stimulated with either calcium ionophore A23187 (Fig. 1A) or TNFα (Fig. S1A) led to significant syndecan-4 release. To test whether soluble factors released from neutrophils are sufficient for glycocalyx degradation, we treated GEnCs with neutrophil conditioned media (CM), containing spontaneously released or induced degranulation contents. Syndecan-4 shedding was triggered by naive neutrophil CM, and the response was further enhanced with CM from A23187-or TNFα treated neutrophils (Fig. 1B). Glycocalyx degradation occurred without significant changes in endothelial cell viability (Fig. S1B).

**Figure 1.**
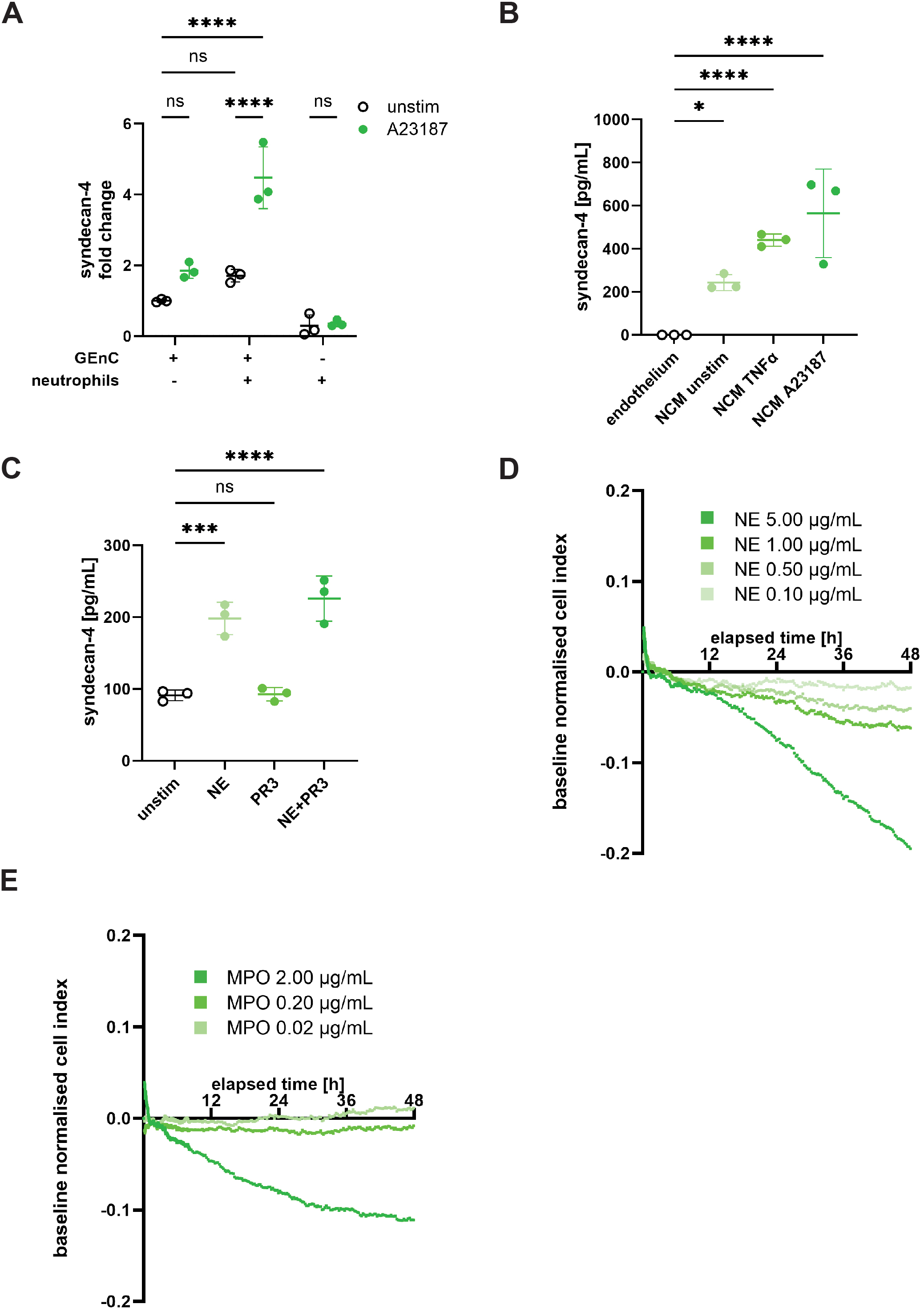
Neutrophil degranulation enhances endothelial permeability and glycocalyx shedding. **A.** Fold change of syndecan-4 in GEnC culture supernatants following co-culture with peripheral blood neutrophils for 5 h, with and without treatment with calcium ionophore A23187 (2.5 μM), n = 3 donors. **B.** Syndecan-4 levels in GEnC culture supernatants following 5 h incubation with neutrophil-conditioned media (NCM) collected after stimulation of neutrophils for 4h with TNFα (2 ng/mL) and A23187 (2.5 μM), n = 3 donors. **C.** Syndecan-4 levels in GEnC culture supernatants following co-incubation with recombinant neutrophil elastase (NE, 1 μg/mL), proteinase 3 (PR3, 1 μg/mL) or both (NE+PR3) for 5 h, n = 3. **D.** TEER cell index of HUVEC cells after treatment with recombinant NE (0.10−5.00 μg/mL) for 48 h, n = 3. **E.** TEER cell index of HUVEC cells after treatment with myeloperoxidase (MPO, 0.02−2.00 μg/mL) for 48 h, n = 3. Data information: Data are presented as mean ± SD. ns – not significant, \**P* ≤ 0.05, \*\*\**P* ≤ 0.001, \*\*\*\**P* ≤ 0.0001, assessed using two-way ANOVA with Tukey’s post-hoc test (**A**) and one-way ANOVA with Dunnett’s post-hoc test (**B-C**).

Neutrophil elastase (NE) and proteinase 3 (PR3) are the most abundant proteases [36] in neutrophil granules and their depletion reduces endothelial dysfunction in mouse models of malaria [17] and sepsis [37, 38]. To test if these proteases can directly degrade the glycocalyx, GEnCs were incubated with recombinant NE and PR3. NE induced significant syndecan-4 shedding, while PR3 had no significant effect (Fig. 1C). Combined NE and PR3 treatment caused only a modest increase in syndecan-4 release, indicating that NE is the principal primary granule protease responsible for endothelial glycocalyx degradation, as previously demonstrated for the secondary granule protease MMP9 [39].

We next tested the effect of primary granule components on endothelial barrier integrity, using the xCELLligence trans-endothelial electrical resistance (TEER) assay, which measures permeability of a cell monolayer. We incubated human umbilical vein endothelial cells (HUVECs) with NE and myeloperoxidase (MPO), two proteins that are strongly and consistently associated with endothelial dysfunction in malaria and sepsis [11, 14, 40, 41]. Both NE and MPO disrupted endothelial barrier integrity in a concentration dependent manner (Fig. 1D-E), at rates similar to the positive control (Dengue virus NS1 protein, known to induce vascular permeability [42]; Fig. S1C-D). In summary, neutrophils can induce endothelial glycocalyx degradation and barrier breakdown, and this is at least partially mediated by degranulation of primary granules.

### Neutrophil proteomic changes associated with severe inflammation

To identify potential targetable mechanisms controlling neutrophil degranulation in systemic infection, we investigated alterations in neutrophil developmental state under severe inflammation. We performed tandem mass tag (TMT) proteomics on neutrophils isolated from peripheral blood of severe malaria and sepsis patients (Fig. 2A). We collected blood from adults hospitalized with *P. falciparum* malaria (n = 6) at Maputo Central Hospital in Mozambique (Table 1) and with septic shock at Bristol Royal Infirmary (United Kingdom, n = 4; Table 2). For each patient group, we collected matched healthy controls (HC) from the same populations. We collected a single admission blood sample from malaria patients, while for sepsis we performed longitudinal sampling at days 1 (T1), 4 (T2), 10 (T3) and 25 (T4) post admission to intensive care (Fig. S2A).

**Figure 2.**
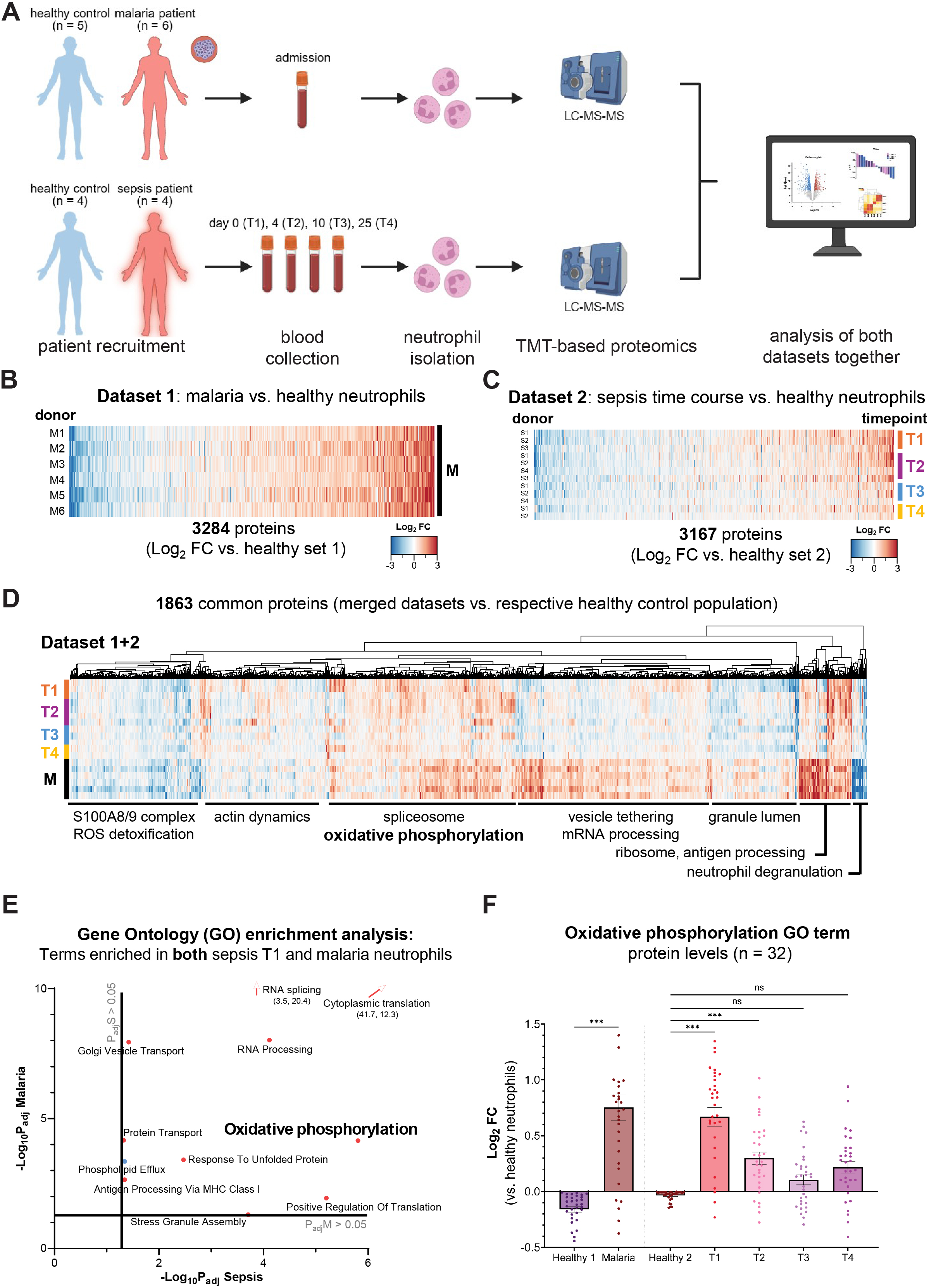
Proteomic analysis of neutrophils from malaria and sepsis patients. **A.** Schematic of proteomics experiment, outlining collection of admission samples from malaria patients and longitudinal sampling of sepsis patients; **B**-**C**. Total dataset overviews of neutrophil protein expression in (B) malaria as compared to HC and (C) sepsis as compared to HC. **D.** Hierarchical clustering (Euclidean distance, complete linkage) of proteins present in both malaria and sepsis datasets, compared to HC samples from the respective datasets. Gene Ontology (GO) enrichment terms for each major cluster are labelled below. **E.** Matching GO terms enriched in separate gene set enrichment analyses of differentially expressed proteins from each dataset. Adjusted *p*-values are mapped in the X-Y plane for each dataset (X: sepsis vs. HC; Y: malaria vs. HC). Terms in red correspond to enrichment results from over-expressed proteins; terms in blue correspond to under-expressed proteins. **F.** Relative abundance of grouped proteins belonging to GO Term ‘*Oxidative phosphorylation* in malaria and sepsis, normalized to respective HCs. Samples from malaria patients were collected at admission; these patients are designated M1–M6. Samples from sepsis patients were collected on days 1, 4, 10 and 25 following admission to the intensive care unit and are designated T1–T4 for each patient (patients S1–S4).

**Table 1:** Clinical charactersitics of malaria patients (Maputo Central Hospital, Mozambique) contributing for neutrophil proteomics analysis.

|  | HC |  | uncomplicated malaria |  | severe malaria |  |
| --- | --- | --- | --- | --- | --- | --- |
| n | 5 |  | 1 |  | 5 |  |
|  | mean (min-max) | % proportion | mean (min-max) | % proportion | mean (min – max) | % proportion |
| Age | 30 (23-36) |  | 36 |  | 34 (22-42) |  |
| Sex (females) | 1 | 20 | 1 | 100 | 2 | 40 |
| Hiv | 0 | 0 | 0 | 100 | 2 | 40 |
| Temperature (C) |  |  | 38.0 |  | 38 (37-40) |  |
| Hb |  |  | 13.2 |  | 12.4 (9.8-12.4) |  |
| WBC |  |  | 3.98 |  | 6.2 (4.2-11.5) |  |
| Neutrophils (%) |  |  | 68% |  | 70% (50-85) |  |
| Platelets |  |  | 25 |  | 48 (19-93) |  |
| Creatinine |  |  | 47 |  | 226<sup>a)</sup> (55-860) |  |
| Glucose |  |  | 4.9 |  | 10.7 (5.7-21.8) |  |
| Respiratory rate |  |  | 16 |  | 22 (16-26) |  |
| Liver failure <sup>b)</sup> |  |  | 0 | 100 | 1 | 20 |
| Coagulat. disturb. <sup>c)</sup> |  |  | 0 | 100 | 1 | 20 |
| Cerebral disturb. <sup>d)</sup> |  |  | 0 | 100 | 1 | 20 |
| Case fatality rate |  |  | 0 | 100 | 0 | 0 |
| Parasitemia <sup>e)</sup> |  |  | 2 |  | 3.4 (3-4) |  |

**Table 2:**
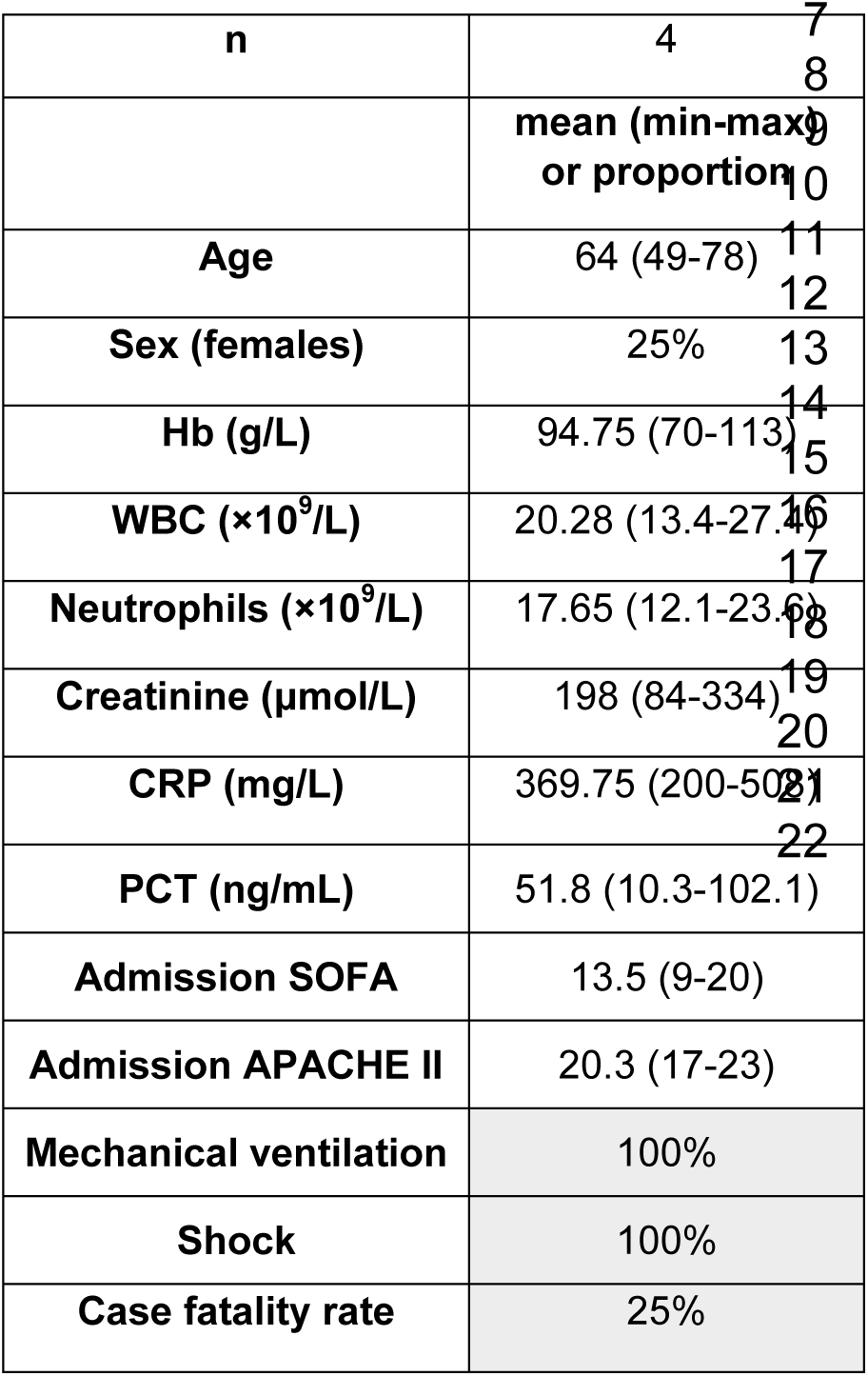
Clinical charactersitics of sepsis patients (Bristol Royal Infirmary, UK) contributing for neutrophil proteomics analysis.

**Table 3:** Cerebral malaria patients (Queen Elizabeth Central Hospital, Blantyre, Malawi), contributing to degranulation analyses.

|  | mean or percentage | min - max |
| --- | --- | --- |
| **Age (in months)** | 50.9 | 17 - 112 |
| **Sex (females)** | 50% |  |
| **Living with HIV** | 10% |  |
| **Temp** | 39 | 36.8 - 39.9 |
| **respiratory rate** | 28 | 24 - 34 |
| **parasitaemia** | 4886 | 406 - 630240 |
| **Hb (g/L)** | 85 | 72 - 102 |
| **WBC ($\times 10^9/L$)** | 8.2 | 4.5 - 15.1 |
| **platelets** | 29.2 | 22 - 51 |
| **Creatinine** | 97.2 | 63.6 -164.0 |
| **Glucose** | 7.2 | 4.3 - 10.3 |
| **Hypoglycaemia** | 0 |  |
| **Liver failure** | 0 |  |
| **Coag disturbances** | 0 |  |
| **CNS disturbances** | 100% |  |
| **Case fatality rate** | 10% |  |

We identified 3284 expressed proteins in the malaria dataset (Fig. 2B) and 3167 in sepsis (Fig. 2C), with 1863 showing differential expression in both diseases (Fig. 2D). The two infections demonstrated some divergences: malaria was exclusively associated with a neutrophil type I interferon response, while sepsis showed elevated activation of neutrophil apoptotic signaling (FigS2B-C). To identify common regulatory pathways in both diseases, we performed gene ontology (GO) enrichment analysis of admission samples. This revealed shared expression patterns of numerous canonical pathways, including upregulation of RNA processing and splicing, as well as cytoplasmic translation (Fig. 2E), indicating strong activation of gene expression in both diseases. Interestingly, neutrophil oxidative phosphorylation (OXPHOS) was among the most significantly upregulated pathways in both diseases (adjusted p-value: malaria=7.09×10^−5^; sepsis=8.09×10^−5^, Fig. 2D-E). Upregulation of 32 OXPHOS proteins was equally prominent in malaria and sepsis admission samples and progressively returned to baseline in later sepsis timepoints; (Fig. 2F). We used the MitoCarta tool [43] to more closely examine expression of mitochondrial proteins: neutrophils from both malaria and sepsis showed elevated expression of mitochondrial central dogma (Fig. S2D), fatty acid oxidation (FAO; Fig. S2E) and calcium homeostasis (Fig. S2F) protein sets, arguing for enrichment of functional mitochondria in patients. Interestingly, TFAM (mitochondrial transcription factor A), a key regulator of mitochondrial DNA maintenance and biogenesis [44], was uniquely upregulated in sepsis but not in malaria (Fig. S2G-H). In summary, proteomic analysis demonstrated enrichment of mitochondria in neutrophils isolated from patients with severe malaria and sepsis, suggesting metabolic rewiring and potential for OXPHOS utilization.

### Increased mitochondrial respiration in infection-elicited neutrophils

We used the Seahorse metabolic flux analyzer to assess infection-induced changes in neutrophil metabolism. Steady-state circulating neutrophils in healthy donors and mice predominantly rely on glycolysis for ATP production and do not engage in OXPHOS [45], however mitochondrial respiration has been detected in mouse neutrophils in cancer models [33]. We initially examined mouse neutrophils from a *Plasmodium chabaudi* (*P. chabaudi*) malaria model. Infection of C57BL/6 mice with the non-lethal *P. chabaudi* clone AS leads to an acute, self-resolving infection with parasitemia peaking at day 9 post-infection (dpi; Fig. S3A). Neutrophils purified from both blood (Fig. 3A) and spleen (Fig. S3B) of infected mice at peak parasitemia (9 dpi) exhibited significantly elevated mitochondrial oxygen consumption rate (OCR) compared to uninfected controls, reflecting increased OXPHOS-driven ATP production (Fig. S3C-D).

**Figure 3.**
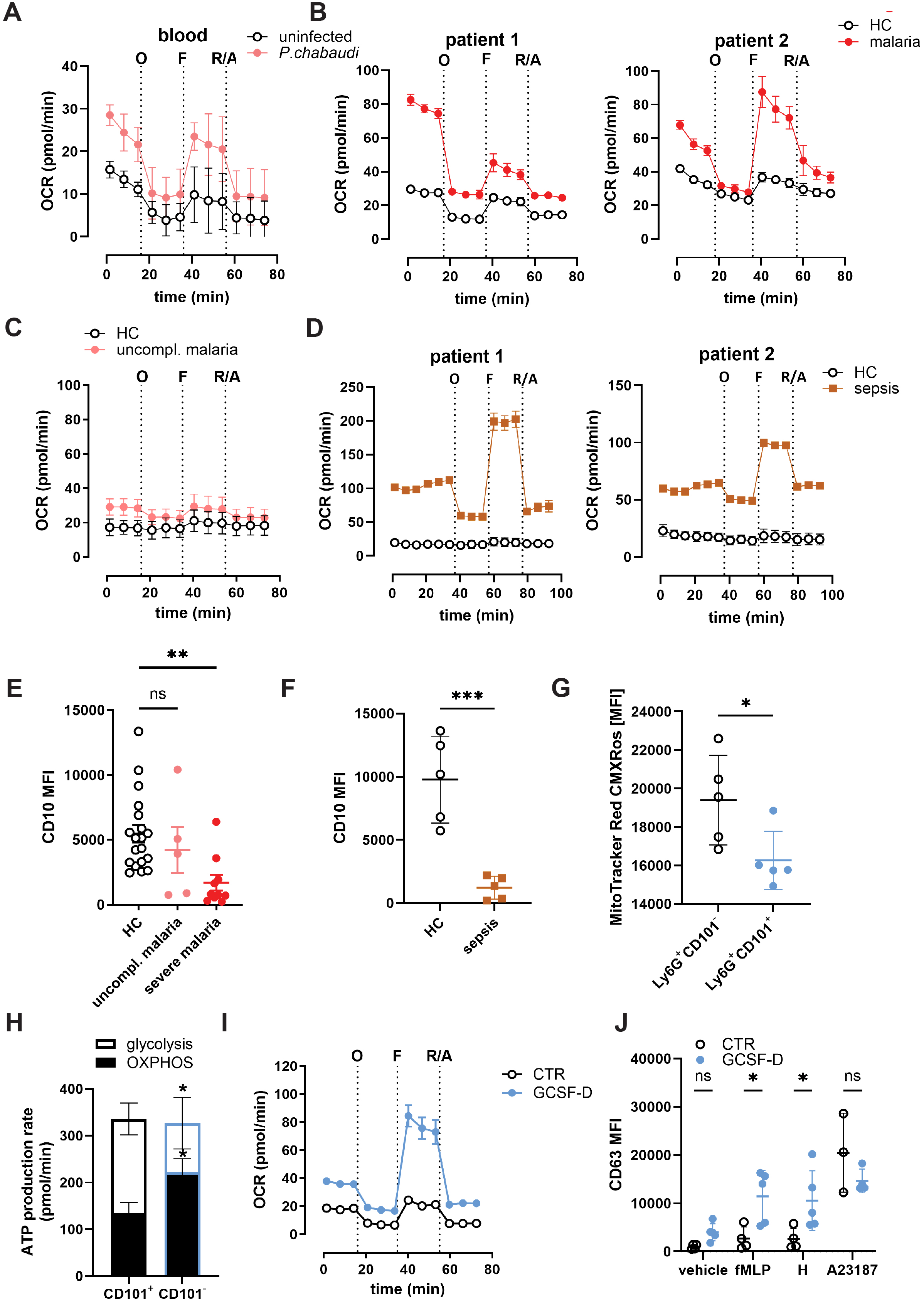
Elevated mitochondrial activity in malaria and sepsis is associated with neutrophil immaturity. **A.** Respiration (OCR) of peripheral blood neutrophils from *P. chabaudi* clone AS-infected mice at 9 dpi (n = 3). **B.** OCR reflecting mitochondrial respiration of neutrophils from HC and two severe malaria patients. **C.** OCR of neutrophils from HC and uncomplicated malaria patient (n = 1). **D.** OCR of neutrophils from HC and two sepsis patients. **E.** Median fluorescence intensity of CD10 on peripheral blood neutrophils from HC (n = 19) and patients with uncomplicated (n = 50 and severe malaria (n = 10). **F.** Median fluorescence intensity of CD10 on peripheral blood neutrophils from HC and sepsis patients (n = 5 each). **G.** Median fluorescence intensity of MitoTracker CMXRos of immature (Ly6G^+^CD101^−^) and mature (Ly6G^+^CD101^+^) neutrophils isolated from bone marrow of uninfected mice, n = 4. **H.** ATP production rate attributed to glycolytic and mitochondrial sources for CD101^−^ and CD101^+^ splenic neutrophils at day 9 post-infection with *P. chabaudi* clone AS, n = 3. **I.** Representative mitochondrial respiration profile of neutrophils from control (CTR) and GCSF-mobilised donors (GCSF-D). **J**. Median fluorescence intensity of CD63 on peripheral blood neutrophils from CTR and GCSF-D stimulated with fMLP (300 nM), heme (30 μM), and A23187 (2.5 μM), n = 4 (CTR), 5 (GCSF-D). Seahorse port injections are indicated on **A-D** and **I** as: O – oligomycin (1.5 μM), F – FCCP (400 nM), R/A – rotenone (1 μM) and antimycin A (1 μM). Data information: Data are presented as mean ± SD. ns – not significant, \**P* ≤ 0.05, \*\**P* ≤ 0.01, \*\*\**P* ≤ 0.001, assessed using one-way ANOVA with Dunn’s post-hoc test (**E**), two-way ANOVA with Šidák’s post-hoc test (**H, J**) and unpaired *t* test (**F, G**).

Next, we examined mitochondrial activity in human neutrophils. As expected, little oxygen consumption was detected in neutrophils from healthy donors (Fig. 3 B and D) and a patient with uncomplicated malaria (Fig. 3C). In contrast, neutrophil mitochondrial oxygen consumption and spare respiratory capacity were strongly elevated in patients with severe malaria (Fig. 3B, n = 2). We observed even higher increases in mitochondrial respiration in neutrophils isolated from two sepsis patients (Fig. 3D). In conclusion, neutrophils elicited by severe systemic infection demonstrate increased capacity and reliance on OXPHOS for ATP production.

### Elevated mitochondrial activity is a feature of immature neutrophils

Systemic inflammation leads to mobilization of immature neutrophils from the bone marrow [28, 46, 47], which have previously been associated with increased mitochondrial metabolism in mouse cancer models [33]. To test whether infection-induced mitochondrial reprogramming correlates with neutrophil immaturity, we used flow cytometry to quantify expression of CD10, a marker that is only expressed on mature cells [47]. Neutrophils from hospitalized malaria patients (Mozambique adults) demonstrated significantly reduced expression of CD10 compared to controls, while patients with uncomplicated malaria showed intermediate expression (Fig. 3E). This was confirmed with a second maturity marker, CXCR2 (Fig. S3E). Similarly, as previously reported [48], septic neutrophils expressed significantly lower CD10 levels at admission, compared to healthy controls (Fig. 3F).

We also evaluated maturity of neutrophils from the blood and spleens of *P. chabaudi*-infected mice, using the mouse maturity marker CD101. We found a striking increase in the proportion of immature (CD101^low^) neutrophils in both circulation (Fig. S3F-G) and the spleen (Fig. S3F and H) during infection with the non-lethal *P. chabaudi* clone AS, as well as with the lethal *P. chabaudi* clone CB (Fig. S3A and S3I).

To assess whether immature neutrophils contain higher mitochondrial mass, we co-stained mouse bone marrow neutrophils with anti-CD101 and MitoTracker CMXRos, a dye that labels metabolically active mitochondria. Immature CD101^low^ neutrophils from naïve mice had significantly increased mitochondrial content compared to CD101^+^ mature neutrophils (Fig. 3G). Next, we sorted splenic neutrophils from *P. chabaudi*-infected mice into CD101^−^ and CD101^+^ populations and analyzed their metabolic profiles using the Seahorse analyzer. ATP production via mitochondrial respiration was elevated in immature neutrophils, whereas mature neutrophils had more active glycolysis (Fig. 3H).

To definitively test whether increased mitochondrial respiration is a general feature of immature human neutrophils, we isolated bone marrow-mobilized neutrophils from healthy stem cell donors treated with granulocyte colony stimulating factor (GCSF).

G-CSF mobilizes both CD34^+^ hematopoietic stem cells and immature bone marrow resident neutrophils [46]. We confirmed a significant reduction in the maturation marker CD10 on peripheral blood neutrophils from GCSF-treated donors (GCSF-D) compared to control donors (CTR); (Fig.S3J-K). Seahorse assays confirmed elevated mitochondrial metabolism in GCSF-D neutrophils (Fig. 3I). This was not simply caused by G-CSF priming, as control (steady state) neutrophils incubated with G-CSF *ex vivo* did not exhibit changes in OCR (Fig. S3L). As expected, immature neutrophils from mobilized blood showed elevated externalization of CD63 (Fig. 3J), demonstrating increased degranulation compared to steady state neutrophils and thereby confirming the link between immature status, elevated mitochondrial activity and degranulation. We conclude that bone marrow mobilization is sufficient for accumulation of neutrophils with active mitochondria and increased degranulation propensity.

### Unique neutrophil mitochondrial dynamics in disease states

To further explore mitochondrial alterations in infection, we analyzed morphology in different neutrophil states. Neutrophils from pediatric malaria cases (n = 4, Blantyre, Malawi), adult sepsis (n=3, Bristol, UK) and GCSF-mobilised donors (n=3, Bristol, UK) were stained with MitoTracker CMXRos and imaged using high-resolution confocal microscopy. Age and sex matched control donors were included for all patients. Mitochondrial abundance did not significantly differ between patients, GCSF-treated donors and controls (Fig. 4A-D), which was confirmed with qPCR analysis of mitochondrial DNA (Fig. S4A). Malaria-elicited neutrophils exhibited striking changes in mitochondrial dynamics, with significantly increased average length (Fig. 4E) and volume (Fig. 4F), indicative of active fusion. Septic neutrophils did not show major differences in mitochondrial volume or length (Fig. 4E-F). The proteomic mitochondrial changes detected in malaria and sepsis thus appear to be due to different mechanisms: elongation and networking of mitochondria in malaria and potentially increased abundance in sepsis. This is consistent with the expression pattern of TFAM, which regulates mitochondrial biogenesis and was specifically upregulated in sepsis (Fig. S2G-H).

**Figure 4.**
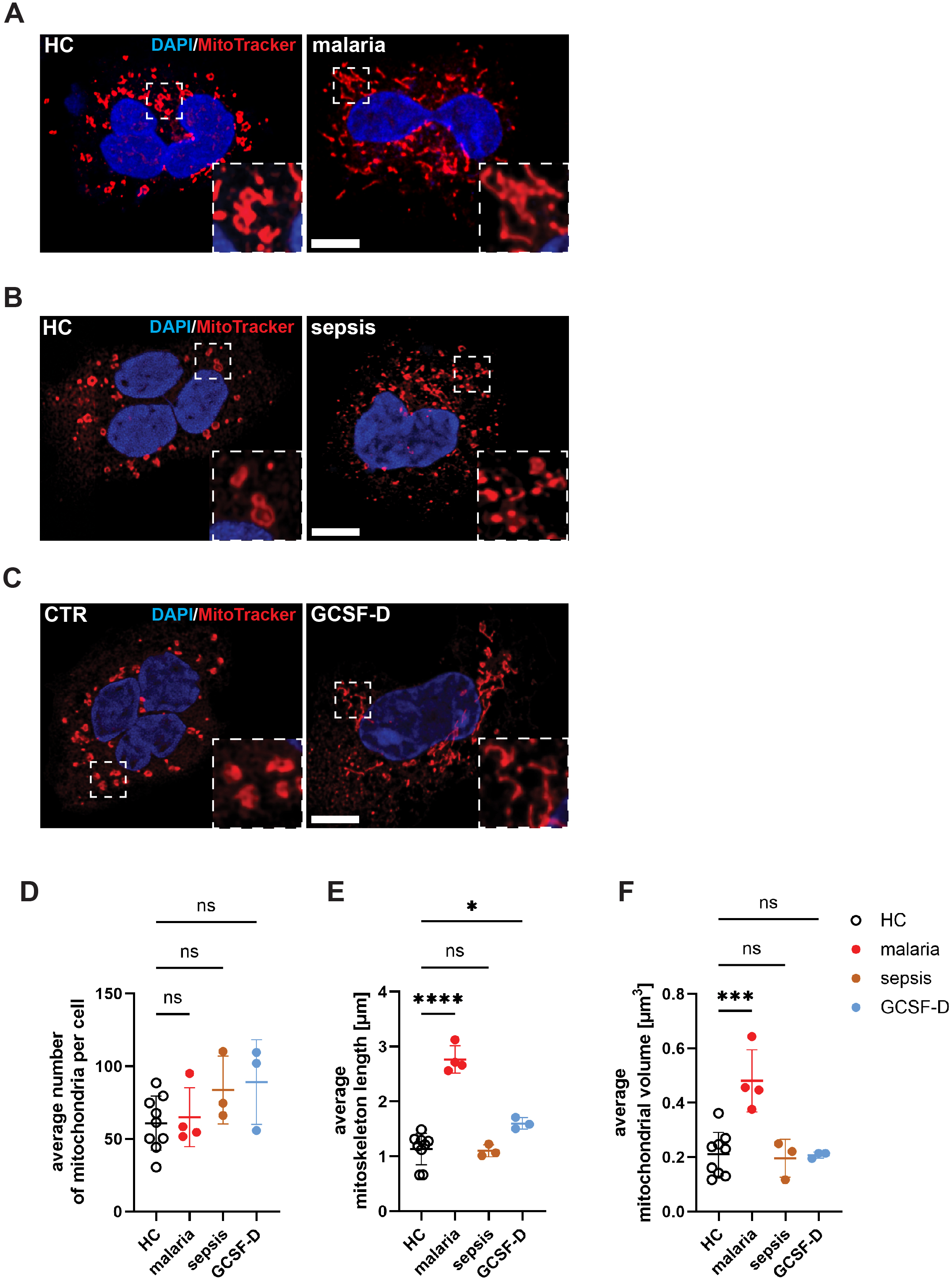
Mitochondrial dynamics differs in malaria– and sepsis-elicited neutrophils. **A-C.** Representative high-resolution confocal images of neutrophils from HC (A), malaria (B) and GCSF-treated donors (GCSF-D, C) stained with MitoTracker CMXRos and DAPI, scale bar 5 μm. **D.** Average number of mitochondria per neutrophil of HC/CTR (n = 9), malaria (n = 4), sepsis (n = 3), and GCSF-D (n = 3). **E.** Average length of mitoskeleton per neutrophil of HC/CTR (n = 9), malaria (n = 4), sepsis (n = 3), and GCSF-D (n = 3). **F.** Average volume of mitochondria per neutrophil of CTR/HC (n = 9), malaria (n = 4), sepsis (n = 3), and GCSF-D (n = 3). Data information: Data are presented as mean ± SD. ns – not significant, \**P* ≤ 0.05, \*\*\**P* ≤ 0.001, \*\*\*\**P* ≤ 0.0001, assessed using one-way ANOVA with Dunnett’s post-hoc test.

### Elevated mitochondrial ROS in inflammation-induced neutrophils

We next tested whether these changes are accompanied by elevated production of mitochondrial ROS (mtROS), an important signaling molecule with immunoregulatory properties [49]. To measure mtROS, we used mitoPY1, a small-molecule fluorescent probe that selectively detects hydrogen peroxide within mitochondria [50]. Circulating neutrophils from naïve mice were MitoPY1-negative, as were bone marrow neutrophils from naïve and *P. chabaudi*-infected mice (Fig. S5A). Interestingly, we detected a substantial population of mtROS-producing neutrophils in the blood of *P. chabaudi*-infected mice, coinciding with peak parasitemia (Fig. 5A, S5A). In parallel, total cellular ROS, assessed by H_2_DCFDA, (primarily reactive with hydroxyl radicals and peroxynitrite), indicated increased total ROS production in neutrophils from both bone marrow and blood of infected mice (Fig. S5A-B). We also observed an increased proportion of MitoPY1^+^ circulating neutrophils in sepsis patients (Fig. 5B) and in GCSF mobilized blood (Fig. 5C), and a trend for increased MitoPY1 fluorescence in pediatric malaria patients at admission, compared to controls (Fig. S5C). We conclude that immature neutrophils with altered mitochondrial structure generate increased mitochondrial hydrogen peroxide compared to steady state.

**Fig. 5.**
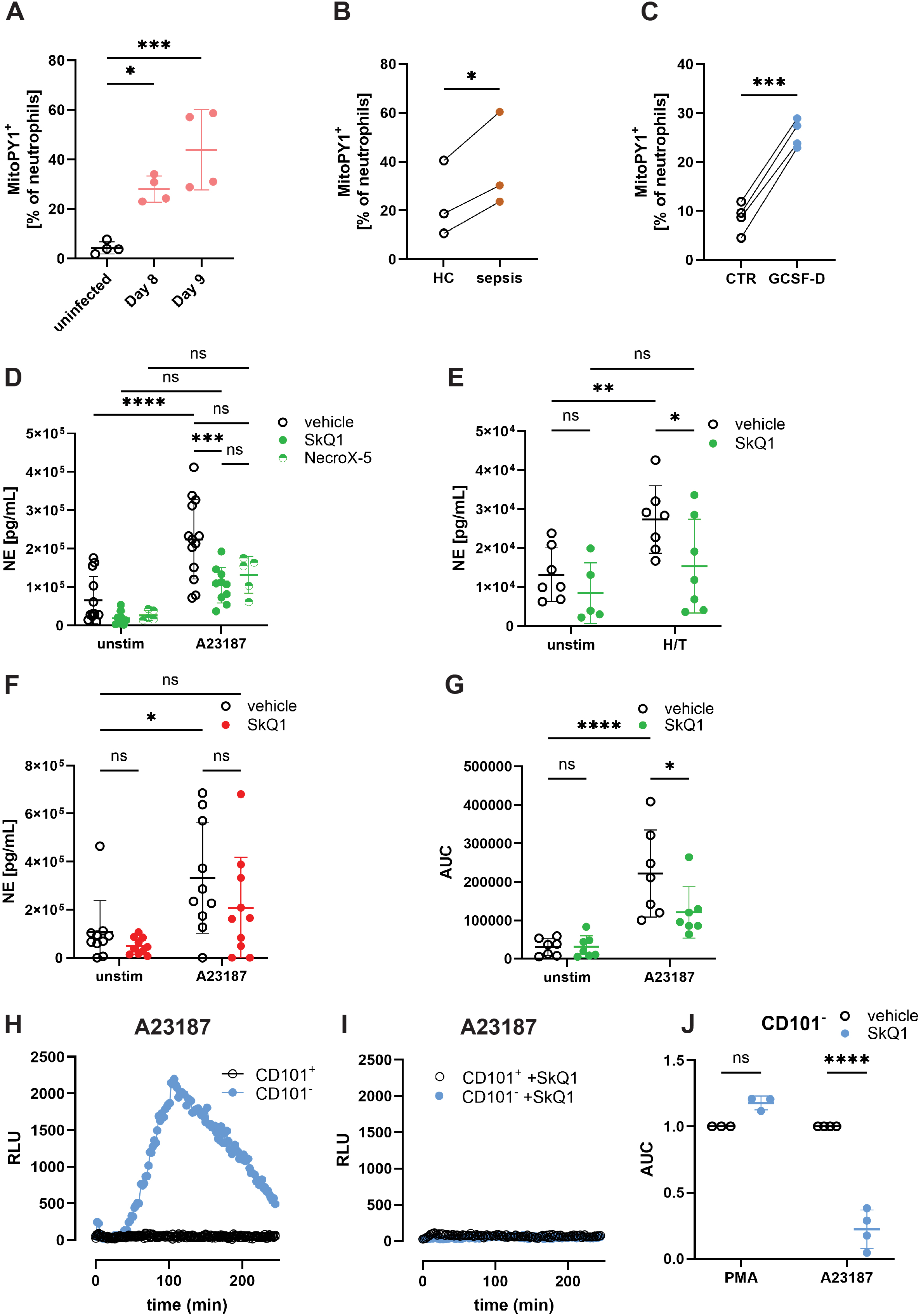
Mitochondrial ROS enhances neutrophil degranulation and oxidative burst. **A.** Percentage of MitoPY1-positive neutrophils from naive and *P. chabaudi* clone AS-infected mice at day 8 and 9 post-infection, n = 4 per group. **B.** Percentage of MitoPY-1 positive neutrophils from HC and sepsis patients, n = 3 each. **C.** Percentage of MitoPY1-postive neutrophils from CTR and GCSF-D donors, n = 4 each. **D.** Neutrophil elastase (NE) levels in supernatants of A23187-stimulated (2.5 uM) CTR neutrophils, pre-incubated with vehicle (DMSO; n = 13), SkQ1 (500 nM; n = 9) or NecroX-5 (5 μM; n = 5). **E.** NE levels in supernatants of CTR neutrophils primed with TNFα (2 ng/mL) and stimulated with hemin (30 μM, H/T), pre-incubated with vehicle (DMSO) or SkQ1 (500 nM), n = 7. **F.** NE levels in supernatants of neutrophils from cerebral malaria patients stimulated with A23187 (2.5 μM), pre-incubated with vehicle (DMSO) or SkQ1 (500 nM), n = 10. **G.** Quantification of oxidative burst (as area under curve, AUC) of CTR neutrophils stimulated with A23187 (5 μM), pre-incubated with vehicle (DMSO) and SkQ1 (500 nM) n = 7. **H.** Representative luminol-based measurement of ROS production in FACS-sorted mouse CD101^−^ and CD101^+^ neutrophils, stimulated with A23187 (5 μM). **I.** Representative luminol-based measurement of ROS production in FACS-sorted mouse CD101^−^ and CD101^+^ neutrophils stimulated with A23187 (5 μM) and pre-stimulated with vehicle or SkQ1 (500 nM), n = 4. **J**. Quantification of oxidative burst (as AUC) of mouse CD101^−^ neutrophils stimulated with PMA (100 nM) or A23187 (5 μM), pre-incubated with vehicle (DMSO) and SkQ1 (500 nM), n = 3. Data information: Data are presented as mean ± SD. ns – not significant, \**P* ≤ 0.05, \*\**P* ≤ 0.01, \*\*\**P* ≤ 0.001, \*\*\*\**P* ≤ 0.0001, assessed using one-way ANOVA with Dunnett’s post-hoc test (**A**), paired *t* test (**B**, **C**) and two-way ANOVA with Tukey’s post-hoc test (**D**-**G**) or Šidák’s post-hoc test (**J**).

### mtROS regulates neutrophil degranulation, oxidative burst and cytokine release

To test whether mtROS can regulate granule exocytosis, we first validated the small molecule SkQ1 [51], a mitochondria-targeted ROS quencher. SkQ1 effectively inhibited neutrophil mtROS *ex vivo* (Fig. S5D). We then pre-incubated neutrophils from healthy donors with SkQ1 and quantified the release of NE and MPO, in response to A23187 and to a combination of TNFα and extracellular heme (H), a malaria and sepsis-relevant stimulus [17, 52]. mtROS quenching significantly reduced primary granule exocytosis in response to both stimuli (Fig. 5D-E, S5E). We confirmed this using a second mtROS scavenger, NecroX-5, which similarly reduced NE release following A23187 stimulation (Fig. 5D). mtROS quenching also impaired release of metalloproteinase-containing secondary granules in response to heme stimulation (Fig. S5F). To validate this mechanism in a disease context, we tested whether mtROS quenching affects degranulation of neutrophils from cerebral malaria patients. SkQ1 treatment reduced NE release by 1.6-fold (HC = 331.7 ng/mL vs. malaria = 207.00 ng/mL; Fig. 5F), although this difference did not reach statistical significance (p = 0.372). SkQ1 stabilized cortical actin (Fig. S6A-B), which needs to be degraded for granules to fuse with the plasma membrane [53], thus providing a potential mechanism for the inhibitory effect on granule exocytosis. Together, these data confirm a regulatory role for mtROS in neutrophil degranulation.

To test whether mtROS regulates other neutrophil functions, we examined the effect of SkQ1 on the NOX2-dependent oxidative burst, using the luminol assay in healthy donor neutrophils stimulated with PMA, a potent PKC agonist, or with A23187. Pre-incubation with SkQ1 had no effect on the PMA oxidative burst (Fig. S5G), but inhibited ROS production with the calcium ionophore (Fig. 5G), suggesting a specific effect on calcium signaling pathways.

To determine whether mitochondrial ROS drive hyper-responsiveness of immature neutrophils to calcium signaling, we first tested differences in calcium-mediated activation. We FACS-sorted naïve mouse bone marrow neutrophils into immature (CD101^−^) and mature (CD101^+^) subsets and analyzed the NOX2-oxidative burst with the luminol assay. The PMA-induced oxidative burst was comparable between the two subsets (Fig. S5H). Interestingly, calcium ionophore A23187 exclusively triggered a robust oxidative burst in immature CD101^−^ but not in CD101+ neutrophils (Fig. 5H), demonstrating that immature neutrophils are more responsive to calcium mobilization. Notably, SkQ1 treatment strongly reduced A23187-induced ROS production in CD101^−^ neutrophils (Fig. 5I-J), whereas PMA-induced ROS remained unaffected (Fig. S5I, 5J).

We also investigated the role of mtROS in neutrophil cytokine and chemokine production. SkQ1 potently suppressed production of IL-8 (Fig. S5J) and IL-6 (Fig. S5K) in response to both LPS (TLR4 ligand) and resiquimod (TLR7/8 ligand).

In summary, our results demonstrate that immature neutrophils in malaria, sepsis and mobilized blood exhibit increased mtROS production, which promotes primary granule exocytosis, NOX2-dependent oxidative burst and inflammatory cytokine production, particularly in response to calcium-dependent stimuli, suggesting an interplay between mtROS production and calcium signaling in regulating neutrophil effector functions.

## DISCUSSION

Fine tuning the neutrophil response in infection involves striking a balance between releasing sufficient antimicrobial granule proteins and limiting host damage. We show that NE and MPO induce endothelial dysfunction by compromising barrier integrity and promoting glycocalyx shedding. This is consistent with reports implicating these proteins in vascular damage [54, 55] and pathogenesis of severe malaria [14, 15, 56] and sepsis [41, 57, 58]. NE and MPO likely synergize with other pro-inflammatory factors to cause vascular dysfunction [59, 60], but have also been individually validated as targets in pre-clinical models of malaria [18] and sepsis [19] and in limited clinical studies [25, 61].

Our study contributes to mounting evidence that mitochondria play a central role in regulating neutrophil activity [30, 33, 62–64]. Importantly, we show that this is particularly relevant in immature neutrophils from infected patients. Immature neutrophils use OXPHOS to generate ATP and higher amounts of mtROS; this is in contrast to neutrophils from healthy donors, which at steady state mainly generate ATP from glycolysis [65]. The striking changes in abundance of mitochondrial proteins in immature neutrophils are consistent with previous omics results showing increased expression of mitochondrial components in neutrophil progenitors [64, 66–68], arguing for a direct link between emergency granulopoiesis and maintenance of OXPHOS activity. In mice, mitochondrial fatty acid oxidation in neutrophil precursors is essential for terminal maturation [69]. We show that in addition to metabolic and developmental functions, mitochondria also regulate calcium-dependent functional responses.

Immature neutrophils have been associated with hyperactive responses [28], particularly in response to calcium mobilization [31]; we show that this is at least partially mediated by enhanced mitochondrial ROS production. In zebrafish infection models, emergency granulopoiesis was shown to upregulate neutrophil oxidative phosphorylation and mtROS [70], which promoted bactericidal activity [70–72]. Taken together with our patient data, this suggests that emergency granulopoiesis augments both quantity and inflammatory capacity of metabolically adapted circulating neutrophils. This may be protective at early stages of infection [70] however enhanced mitochondrial activity also contributes to neutrophil hyperactivity and ensuing immunopathology.

MtROS are key signaling molecules in innate immunity and are known to activate neutrophil [73–75] and macrophage responses [76]. Mechanistically, mtROS can promote stabilization of the NF-κB activator NEMO, which induces production of pro-inflammatory cytokines by macrophages [77] and can activate MAP kinase signaling [78]. In neutrophils, mtROS have been shown to directly kill bacteria [70, 79, 80], as well as extend lifespan by stabilizing the pro-survival factor Hif1α [81]. In other cells, mtROS has been shown to regulate cytosolic calcium concentrations [82], providing a potential mechanistic explanation for the specific effects we observed with calcium ionophore.

MtROS quenching has been shown to have anti-inflammatory effects *in vivo*: SkQ1 had significant protective effects in a mouse model of sepsis [83], while mitoTEMPO was shown to reduce neutrophil activation and pro-inflammatory cytokine production in influenza [84] and lupus [30]. Interestingly, a mutation in *SOD2*, encoding an enzyme that promotes breakdown of mtROS, has been linked to malaria severity in a genome-wide association study [85].

We observed differences in mitochondrial dynamics between malaria and sepsis. Although this requires additional investigation, neutrophils from malaria patients contain mitochondria of greater length and volume, suggesting active fusion mechanisms. Neutrophils from sepsis patients, on the other hand, may be engaging mitochondrial biogenesis, as shown by increases in mitochondrial chromosome copy number and TFAM expression. Both fusion and biogenesis are responses to mitochondrial stress and may be reflective of differing inflammatory milieus or glucose levels in the two diseases [86]. Sepsis is generally associated with higher levels of inflammation, glucose and emergency granulopoiesis than malaria [14, 15, 87] and it would be interesting to correlate these parameters with mitochondrial morphology and neutrophil activity.

### Limitations of our study

Although we tested a variety of clinical samples, the sample size was often low, reflecting the major challenges with performing neutrophil assays in real-time on fresh samples in malaria endemic settings. Malaria and sepsis are both heterogenous conditions and breakdown into disease subtypes would add granularity to our findings, but this would require a much larger study. Furthermore, due to restrictions to patient blood access, some of our functional studies with SkQ1 were performed on neutrophils from healthy donors, which contain fewer active mitochondria. All mtROS scavenging should be repeated with patient samples, rather than steady state neutrophils; we anticipate stronger effect size in immature metabolically adapted neutrophils. Although we used multiple mtROS scavenging compounds, inhibitors are likely to have off-target effect and our conclusions would benefit from genetic validation, such as modulation of mtROS scavenging enzymes.

Many questions remain before neutrophil mitochondria can be targeted for therapeutic purposes. Is signaling predominantly mediated by oxidation of protein targets, as described for NEMO [77], or do other pathways such as altered cytosolic calcium mediate the downstream effects of mtROS signaling? How does mtROS affect neutrophil development and heterogeneity?

Loss of mitochondrial homeostasis has been proposed as a unifying principle in chronic and acute inflammation [88]. Neutrophils, long thought to have negligible and inactive mitochondrial networks, are emerging as potential targets for mitochondrial drugs, which may be a promising avenue for reversing endothelial damage in diseases such as malaria and sepsis.

## MATERIALS AND METHODS

### Blood collection and patient recruitment

Written informed consent was obtained from all patients and healthy donors, or from guardians if patients were too unwell to consent. Sample collection at Maputo Central Hospital was approved by the National Ethics Committee in Mozambique (594/CNBS/2018) and by the Regional Ethics Committee in Southeastern Norway (2015/2097/REK South-East A). Sample collection in Malawi was approved by the College of Medicine Research Ethics Committee, University of Malawi (now Kamuzu University of Healthy Sciences) [13/12/2019, P.11/19/2866 Immunopathogenesis of Cerebral Malaria (IMPAC)], by the Liverpool School of Tropical Medicine Research Ethics Committee (07/02/2020 Research protocol 19-099) and by the Medical, Veterinary and Life Sciences Research Ethics committee at the University of Glasgow (09/03/2020 Research protocol 200190088).

Sepsis samples were obtained through the Immune Cell metabolism in Intensive Care (METABOLIC) study in the Bristol Royal Infirmary, Bristol, UK. This study was approved by NHS REC 22/LO/0678.

For Seahorse assays, patients with imported *P. falciparum* malaria were recruited from hospitals in London, UK as part of the SCRIPT study (ClinicalTrials.gov ID NCT05149157) under ethical approval of Northeast Newcastle & North Tyneside 1 Research Ethics Committee (22/NE/0005).

Collection of blood from healthy donors at the University of Bristol was approved by NHS REC 18/EE/0265.

### Mice and murine malaria model

All procedures were performed under UK home office project license PPL PF2C617B4. Male and female C57BL/6J mice (aged 7-15 weeks) were purchased from Charles River. To initiate *P. chabaudi* infection, experimental mice were infected by intraperitoneal injection with 1 × 10^5^ iRBCs in PBS. Age– and sex-matched uninfected mice were used as controls. Mice were routinely monitored for adverse effects, and supportive treatment, such as wet mash feed and heat pads, were provided to minimize adverse effects. At peak parasitemia (9 dpi), mice were sacrificed by cardiac puncture exsanguination under terminal anesthesia, and blood and plasma were collected for analyses.

### Neutrophil isolation

Human neutrophils were isolated from whole venous blood collected with BD Vacutainer® K2EDTA or BD Vacutainer 3.2% Sodium Citrate tubes using EasySep^TM^ direct human neutrophil isolation kit (STEMCELL Technologies, Cambridge UK) as per manufacturer’s instructions.

Mouse neutrophils were isolated from either bone marrow, spleen or blood. Bone marrow was extracted by flushing tibias and femurs with cold sterile PBS, and then extracted cells were subjected to red blood cell lysis using ACK lysis buffer (Gibco). Spleens were homogenized using a syringe plunger and a 70 µm cell strainer. Cell strainers were washed with ice-cold PBS, and resultant cell pellet was washed with ACK lysis buffer and lysate was refiltered through a 70 µm cell strainer. Blood was extracted by cardiac puncture under terminal anesthesia and extracted into heparin (100 U/mL), and then red blood cell lysis was performed by using ACK lysis buffer. All cell suspensions were incubated with Fc block (anti CD16/32 Fc block, 4 μg/mL, clone 2.4G2, BD Biosciences) for 5 min on ice, followed by incubation with anti-Ly6G biotin (20 μg/mL, clone IA8, BioLegend) for 30 min on ice. Cells were washed and incubated with streptavidin beads (Miltenyi Biotec) for 15 min at 4 °C. Cells were washed and then added to a LS column (Miltenyi Biotec) as per manufacturer’s instructions.

### GEnC cultures, stimulations and glycocalyx component detection

Kidney endothelial cells (GEnC) were cultured as previously described [35]. Briefly, GEnCs were cultured in Endothelial Cell Growth Basal Medium 2 (EBM-2, PromoCell) containing 5% fetal bovine serum (FBS), 0.04% hydrocortisone, 0.4% human fibroblast growth factor (hFBF-B), 0.1% vascular endothelial growth factor (VEGF), 0.1% R3 insulin like growth factor (R3 IGF-1), 0.1% ascorbic acid, 0.1% human epidermal growth factor (hEGF), and 0.1% GA-1000 at 33 °C with 5% CO_2_. Once 80% confluence was reached, cells were transferred to 37 °C with 5% CO_2_ and cultured for an additional 7 days to mature the GEnC glycocalyx.

GEnCs were stimulated with neutrophil elastase (1 μg/mL), proteinase 3 (1 μg/mL) or purified neutrophils (3 × 10^6^/mL) for 5 h at 37 °C with 5% CO_2_. Neutrophils were additionally stimulated with TNFα (2 ng/mL) or calcium ionophore A23187 (2.5μM). Supernatants were removed and centrifuged to remove cellular debris and a human syndecan-4 (hSDC4) ELISA (DuoSet, R&D Systems) was performed as per manufacturer’s instructions.

To assess cell death in co-cultures, neutrophils and GEnC cells were detached using accutase and centrifuged at 350 × g for 5 min. Cells were resuspended in 100 μL viability dye (Zombie Aqua^TM^ at 1:1000 in PBS) and incubated for 10 min at RT in the dark. Cells were washed twice with PBS and fixed in 4% PFA for 20 min in the dark, followed by a final centrifugation and resuspension in PBS. Samples were acquired using a BD LSRFortessa^TM^ X-20 flow cytometer and data were analyzed using FlowJo (FlowJo, LLC).

### HUVEC culture and trans-endothelial electrical resistance (TEER) assay

Human umbilical vein endothelial cells (HUVECs) from pooled donors were sourced from Merck (Sigma-Aldrich, C-12203). They were cultured in Endothelial Cell Growth Medium 2 (EGM-2, Sigma-Aldrich) and culture vessels pretreated with fibronectin (1 µg/mL, Sigma-Aldrich). The cells were not used beyond passage 5.

An xCELLigence Real Time Cell Analyzer (Agilent) was used to measure the integrity of a HUVEC endothelial layer after treatment with recombinant proteins. Cells were seeded according to manufacturer’s recommendations into a PET E-plate (Agilent) that was pretreated with fibronectin (1 µg/mL) at 30,000 cells per well and allowed to come to confluency for 18 hours. Cell Index values were checked prior to proceeding to ensure suitable confluency had been reached in all wells. Recombinant proteins were prepared in 2× concentrations in EGM-2 and pre-warmed in a 5% CO2 incubator for 30 min. To minimize disruption to the cells during addition of treatment, 100 µL of media per well was removed and 100 µL of 2× treatment media or fresh media were replaced rapidly. Final concentrations were: myeloperoxidase (MPO, Sigma-Aldrich) at 2 µg/mL, 200 ng/mL and 2 ng/mL; neutrophil Elastase (NE, Enzo) at 5 µg/mL, 1 µg/mL, 0.5 µg/mL and 100 ng/mL; and Dengue virus serotype 2 NS1 protein (The Native Antigen Company) at 5 μg/mL. Measurements were recorded every 30 s for 8 h and then every 15 min for 40 h.

### Fluorescence-activated cell sorting (FACS)

CD101^+/-^ mouse neutrophil populations were isolated using FACS following extraction as described above. Purified neutrophil populations were incubated in staining buffer (PBS with 0.5% BSA and 5 mM EDTA) containing Fc block (anti CD16/32 Fc block, BD Biosciences) for 5 min on ice, following which cells were stained with anti-Ly6G BV421 (IA8, BioLegend) and anti-CD101 PE (Moushi101, Invitrogen) for 20 min on ice. Cells were washed and resuspended in staining buffer containing DRAQ7 (Biostatus). Gating eliminated doublets and DRAQ7^+^ dead cells. Fluorescence minus one (FMO) controls were used to set CD101^+/-^ gates. Cells were sorted using a BD Aria II sorter with 70 μm nozzle and into chilled, sterile, heat-inactivated FBS.

### Mitochondrial network analysis by super-resolution microscopy

500 μL of RPMI containing MitoTracker CMXRos (200 nM, Invitrogen) was added to 24-well plates containing 13 mm coverslips (VWR) pre-sterilized with 100% ethanol. 5 × 10^4^ purified neutrophils were resuspended in 50 μL RPMI and added directly to the media-containing wells. Cells were incubated at 37°C, 5% CO_2_ for 20 min. Cells were fixed by adding 500 μL 4% paraformaldehyde (PFA) directly to the medium (final concentration 2% PFA) and incubated for 30 min at 37 °C. PFA-containing media was removed, and coverslips were washed twice with fresh PBS. Coverslips were mounted in ProLong^TM^ Gold Antifade Mountant with DNA stain DAPI (Invitrogen). Super-resolution images of mitochondria were acquired using a Leica SP8 AOBS confocal laser scanning microscope with 63×/1.4NA oil objective with 1.3× pre-acquisition zoom, followed by LIGHTNING adaptive image restoration. To quantify mitochondrial number and mitoskeleton length and volume, images were analyzed using the Fiji Modular Image Analysis (MIA) plugin [89] with a custom workflow. At least 40 cells were imaged and analyzed per conditions across three independent experiments.

### mtDNA quantification using qPCR

2 × 10^6^ purified neutrophils were pelleted, and DNA was extracted using QIAamp® DNA Micro kit (Qiagen) as per manufacturer’s instructions. DNA was quantified using a NanoDrop and 25 ng DNA was used to assess application of mtDNA and nuclear DNA (gDNA) using previously published primers [90] with SYBR^TM^ Green Master Mix (Applied Biosystems) on a QuantStudio^TM^ 3 (Applied Biosystems). Cycling conditions were as follows: step 1 – 1× 95 °C for 20 s, step 2 – 40× 95 °C for 3 s and 56 °C for 30 s. Fold change in mtDNA was assessed as in [90].

### Flow cytometry staining

Staining of whole blood neutrophils from mouse and human was performed as follows. 100 µL of whole blood or purified neutrophils were centrifuged at 350 × g, and the pellet was resuspended in Fc block (human: FcX^TM^ TruStain, BioLegend; mouse: anti-CD16/32 Fc block, BD Biosciences) in staining buffer (PBS, 0.5% BSA and 5 mM EDTA) and incubated on ice for 5 min. Primary antibodies in staining buffer were added directly to Fc-blocked cells and incubated in the dark on ice for 20 min. Next, in case of whole blood samples, cells were washed three times in ACK lysis buffer to remove excess red blood cells, and then all cell samples were fixed in 4% PFA for 20 min in the dark at RT. Samples were acquired using a BD LSRFortessa^TM^ X-20 flow cytometer, and data were analyzed using FlowJo^TM^ (FlowJo, LLC). The list of antibodies used is provided in the table below.

| Antibody | Source | Catalog number |
| --- | --- | --- |
| PE anti-human CD10 | BioLegend | 312204 |
| Brilliant Violet 421<sup>TM</sup> anti-human CD63 | BioLegend | 353030 |
| APC anti-human CD66b | BioLegend | 305118 |
| PerCP/Cyanine5.5 anti-human CD101 | BioLegend | 331016 |
| PE/Dazzle<sup>TM</sup> 594 anti-human CD182 (CXCR2) | BioLegend | 320722 |
| CD101 (Moushi101), Alexa Fluor<sup>TM</sup> 700 | Invitrogen | 56-1011-82 |
| Brilliant Violet 421<sup>TM</sup> anti-mouse Ly-6G | Biolegend | 127628 |

### mtROS and total ROS

0.5 × 10^6^ mouse bone marrow cells, ACK-lysed mouse whole blood, or 0.5 × 10^6^ purified human neutrophils were resuspended in 100 µL of HBSS with Ca^2+^/Mg^2+^ and incubated for 15 min at 37 °C with 5% CO_2_. Next, 100 µL of pre-warmed HBSS+Ca^2+^/Mg^2+^ containing either MitoPY1 or H_2_DCFDA was added to reach a final concentration of 10 µM (MitoPY1) or 5 µM (H_2_DCFDA), followed by a 20-minute incubation at 37 °C with 5% CO_2_. For mouse bone marrow and whole blood samples, following incubation, cells were then washed with pre-warmed HBSS and resuspended in staining buffer (PBS with 0.5% BSA and 5 mM EDTA) containing Fc block (anti-CD16/32, BD Biosciences). Samples were incubated on ice in the dark for 5 min, and then stained with anti-Ly6G BV421 antibody (BioLegend) for 20 min. After antibody staining, or directly after MitoPY1 staining in the case of purified human neutrophils, cells were washed with staining buffer, and events were acquired using an Agilent ACEA NovoCyte 3000 Flow Cytometer, followed by analysis with FlowJo software (FlowJo, LLC).

For live-imaging of MitoPY1-stained neutrophils, cells were resuspended in 500 µL of HBSS with Ca^2+^/Mg^2+^ supplemented with either DMSO or SkQ1 (500 nM), seeded into chambered µ-Slide coverslip (Ibidi) and incubated for 15 min at 37 °C with 5% CO_2_ to allow cell attachment. Subsequently, 500 µL of pre-warmed HBSS+Ca^2+^/Mg^2+^ containing MitoPY1 was added to reach a final concentration of 10 µM, followed by a 20-minute incubation at 37 °C with 5% CO_2_. After incubation, cells were imaged using a Leica DMI6000 inverted epifluorescence microscope, and images were analyzed using Fiji (ImageJ2). At least 40 cells per condition were imaged and analyzed across three independent experiments.

### Degranulation

5 × 10^5^ neutrophils were resuspended in 100 µL RPMI supplemented with HEPES and 0.025% HSA and incubated at 37 °C with 5% CO_2_ for 15 min. For mtROS quenching experiments, cells were pre-incubated in media containing either SkQ1 (500 nM) or NecroX-5 (5 µM) for 30 min at 37 °C with 5% CO_2_. After incubation, cells were stimulated with hemin (30 µM, preceded by 15 min of priming with 2 ng/mL TNFα, Frontier Scientific) or calcium ionophore A23187 (2.5 µM, Sigma-Aldrich) for 60 min at 37 °C. Following stimulation, cells were centrifuged at 350 × g for 5 min at 4°C, and supernatants were collected for further analysis.

For degranulation analysis of GCSF-D neutrophils, cells were stimulated with fMLP (300 nM, Sigma-Aldrich), hemin (30 µM) or A23187 (2.5 µM) for 30 min at 37 °C. Following stimulation, cells were stained and analyzed according to flow cytometry staining protocol described above.

### Cytokine release assays

1 × 10^5^ neutrophils were resuspended in 200 µL RPMI supplemented with 10% FBS and P/S and incubated at 37 °C with 5% CO_2_ for 45 min. After incubation, cells were stimulated with *E. coli* LPS (100 ng/mL; Sigma-Aldrich) or resiquimod (R-848, 5 µM; Sigma-Aldrich) overnight at 37 °C. Following stimulation supernatants were collected for further analysis.

### Detection of soluble neutrophil proteases and cytokines

Supernatants obtained after neutrophil degranulation and cytokine release assays were analyzed using Human Myeloperoxidase DuoSet ELISA (R&D Systems), Human Neutrophil Elastaste/ELA2 DuoSet ELISA (R&D Systems), Human IL-6 DuoSet ELISA (R&D Systems), Human IL-8/CXCL8 DuoSet ELISA (R&D Systems) according to manufacturer’s instructions.

### Seahorse extracellular flux analysis

Seahorse XFe96 sensor cartridge was calibrated overnight as per manufacturer’s instructions (Agilent). Neutrophils were analyzed directly after purification or following the pre-treatment with compounds (see details in figure legends). 4 × 10^5^ cells were resuspended in 180 µL Seahorse XF DMEM supplemented with 5 mM glucose and 2 mM glutamine and plated in a Seahorse XFe96 cell culture microplate. ATP rate assays and mitochondrial stress tests were performed as per manufacturers’ instructions. Final compound concentrations were as follows: oligomycin (1.5 µM), FCCP (500 nM), antimycin A (1 µM), and rotenone (1 µM). Where cell seeding inconsistency was suspected based on variations in post-rotenone and antimycin A oxygen consumption rates (OCR), protein normalization was performed using the Pierce BCA Protein Assay Kit (Thermo Fisher Scientific), and subsequent data were expressed as OCR (pmol/min/µg of protein). Malaria samples for Seahorse analysis were collected in London as part of the SCRIPT study, while sepsis samples were recruited in Bristol as part of the METABOLIC study.

### Measurement of oxidative burst by luminol-enhanced chemiluminescence

1 × 10^5^ neutrophils were resuspended in 100 µL HBSS+Ca^2+^/Mg^2+^ supplemented with HEPES and 0.025% human serum albumin (Griffols). Cells were plated in a white 96-well plate and cultured at 37°C with 5% CO_2_ for 15 min. Horseradish peroxidase (HRP, 1200 U/mL) and luminol (50 mM) were then added, followed by further incubation for 15 min at 37 °C with 5% CO_2_. Neutrophils were subsequently stimulated with calcium ionophore A23187 (2.5 µM) or PMA (50 nM), and chemiluminescence was recorded every 2 min for 3 h using a BMG Labtech FLUOstar Omega plate reader.

### Visualization of F-actin using fluorescently labeled phalloidin

500 μL of RPMI was added to 24-well plates containing 13 mm coverslips (VWR) pre-sterilized with 100% ethanol. 5 × 10^4^ purified neutrophils added directly to the media-containing wells. Cells were incubated at 37 °C, 5% CO_2_ for 15 min to allow attachment after which vehicle (DMSO) or SkQ1 (MedChemExpress) was added to a final concentration of 500 nM. Cells were incubated for 45 min and then fixed for 10 min at 37 °C with freshly prepared 4% methanol-free PFA in PEM buffer (80 mM PIPES, 5 mM EGTA, 2 mM MgCl_2_, pH 6.8). The PFA-containing medium was removed, and coverslips were washed once with PEM buffer. To visualize F-actin, cells were first permeabilized for 10 min with 0.1% Triton-X (in PEM buffer), blocked with 1% BSA (in PEM buffer), and then incubated for 30 min with 150 nM phalloidin conjugated to Alexa Fluor^TM^ 488 (Invitrogen). After washing, coverslips were mounted in ProLong^TM^ Gold Antifade Mountant with DNA stain DAPI (Invitrogen).

Super-resolution images of F-actin cytoskeleton were acquired using a Leica SP8 AOBS confocal laser scanning microscope with 63×/1.4NA oil objective with 1.3× pre-acquisition zoom, followed by LIGHTNING adaptive image restoration. To quantify peripheral actin density in the outer 10% region of the cells, images were analyzed using the Fiji (ImageJ2). In brief, images were projected using Z-stack sum-slices projection, single cells were separated using the polygonal selection tool to measure integrated density of the whole cell, and then selection was scaled to 90%. The difference between the integrated densities of the whole cell and the 90% area represented the integrated density of outer 10% rim. At least 40 cells were analyzed per condition.

### Proteomics

5 × 10^6^ purified peripheral blood neutrophils were washed in PBS and resuspended in RIPA buffer with 10 mM EDTA, 2 mM PMSF (Sigma-Aldrich), 1× Halt Phosphatase (Thermo Fisher Scientific), 50 mM TCEP (Sigma-Aldrich) and Protease Inhibitor Cocktail (Calbiochem). Isolated protein was quantified using the Pierce BCA Protein Assay Kit (Thermo Fisher Scientific) and 100 µg of protein was used for proteomic analysis. Tandem mass tag (TMT) proteomics was performed as described previously [28].

### Bioinformatics

Raw mass spectrometry data were analyzed using Proteome Discoverer (PD) software v. 2 and cross-referenced against the human UniProt database. PD analysis was conducted for full trypsin digestion, removing all hits with more than one missed cleavage. All peptides were filtered to meet an FDR of 1%. Log_2_ fold changes (Log_2_FC) were calculated between neutrophils of 1) malaria patients vs. HC and 2) sepsis patients vs. HC, resulting in quantification of 3284 and 3167 proteins, respectively. Both datasets were merged to create a unified dataset quantifying 1863 proteins. Clustering analysis and the resulting heatmap visualizations were generated using R v. 4.2.2. Gene Ontology (GO) enrichment analyses were performed using the 2025 GO modules within the Enrichr suite [91]. MitoCarta enrichment analyses were performed using MitoCarta 3.0 modules [43] for analysis with the Gene Set Enrichment Analysis tool [92, 93].

### Statistical analysis

Data are presented as mean ± SD. Unless stated otherwise, “n” refers to the number of biological repeats. Statistical analysis was performed using GraphPad Prism 10 software. When applicable, data normality was assessed using the Shapiro-Wilk test prior to further analysis. For comparisons between two groups, an unpaired two-tailed *t* test was used. For comparisons involving more than two groups, one-way or two-way ANOVA followed by post hoc test was applied. Asterisks in graphs indicate significance levels as follows: * *p* < 0.05, ** *p* < 0.01, *** *p* < 0.001 and **** *p* < 0.0001. The absence of asterisks denotes non-significant differences.

## DATA AVAILABILITY

The mass spectrometry proteomics data will be deposited to the ProteomeXchange Consortium at a later date.

The research materials supporting this publication can be accessed by contacting corresponding authors.

## AUTHOR CONTRIBUTIONS

**Przemysław Zakrzewski**: Formal analysis; Investigation; Visualization; Writing— original draft; Writing—review and editing.

**Christopher M Rice**: Formal analysis; Investigation; Writing—original draft.

**Claire Naveh**: Formal analysis; Investigation; Writing—original draft.

**Isaac Dowell**: Formal analysis; Investigation.

**Kathryn Fleming**: Formal analysis; Investigation.

**Aravind V Ramesh**: Resources, Formal analysis.

**Rachel Jones**: Formal analysis; Investigation.

**Pedro L. Moura**: Formal analysis; Investigation.

**Drinalda Cela**: Formal analysis; Investigation.

**Sarah Groves**: Formal analysis; Investigation.

**Stephanie Fletcher-Jones**: Formal analysis; Investigation.

**Yohance Victory**: Formal analysis; Investigation

**Mainga Bhima**: Formal analysis; Investigation

**Stefan Ebmeier**: Sample collection

**Laura Carey**: sample collection, reviewing

**Matthew Buttler**: formal investigation, conceptualisation

**Simon C. Satchell**: conceptualisation

**Ase Berg**: conceptualization, sample collection, formal investigation

**Nadia Palolite:** sample collection, formal investigation

**James Nyirenda**: sample collection, formal investigation

**Watipenge Nyasulu**: sample collection, formal investigation

**Isabel Zgambo**: sample collection, formal investigation

**Charalampos Attipa**: conceptualization, sample collection, formal investigation

**Linda Woolridge**: sample collection, supervision

**Andrew Davidson**: conceptualization, sample collection, formal investigation

**Aubrey Cunnington**: conceptualisation, sample collection, supervision

**Christopher A. Moxon**: conceptualization, formal investigation, supervision, funding, writing (reviewing and editing).

**Borko Amulic**: Conceptualization; Formal analysis; supervision; funding acquisition; writing—original draft; writing, reviewing and editing.

## ACKNOWLEDGEMENTS

We thank all the patients and blood donors for participating in our study, as well as nurses and clinical staff at Maputo Central Hospital (Mozambique), Blantyre Queen Elizabeth Hospital (Malawi), Bristol Royal Infirmary (UK). We acknowledge the assistance and contribution from SCRIPT investigators Dr Aula Abbara, Dr Ana Garcia Mingo, Alicia Chevalier, Abiola Harrison, Dr Maggie Nyirenda, Blessing Kazooba. We acknowledge the help and support of Dr. Karl Seydel, Prof. Terrie Taylor and other investigators, clinicians and nurses on the paediatric research ward, Queen Elizabeth Central Hospital, Malawi.

BA’s lab is funded by the Medical Research Council (MRC), grant MR/R02149X/1. PLM is supported by the Myelodysplastic Syndromes Foundation, Inc. (grant 1142079), the Dr. Åke Olsson foundation (Dnr 2024-00303), the Alex and Eva Wallström foundation (Dnr 2024-00311) and a KI Research Foundation grant (Dnr 2024-02330). AR is supported by the Wellcome Trust through a GW4-CAT-HP fellowship (225540/Z/22/Z). SE and the SCRIPT study were funded by the Imperial 4i Clinician Scientist Training Programme (Lee Family Faculty of Medicine Scholarship Endowment). CAM’s lab is supported by a UKRI MRC Future Leaders Fellowship MR/V025856/1. Malawi-Liverpool-Wellcome Research Programme is supported by core funding from Wellcome. Infrastructure support for this research was provided by the NIHR Imperial Biomedical Research Centre.

The views expressed are those of the authors and not necessarily of the NHS. The schematic summary figures were created in BioRender.

## FIGURE LEGENDS

**Figure S1.**
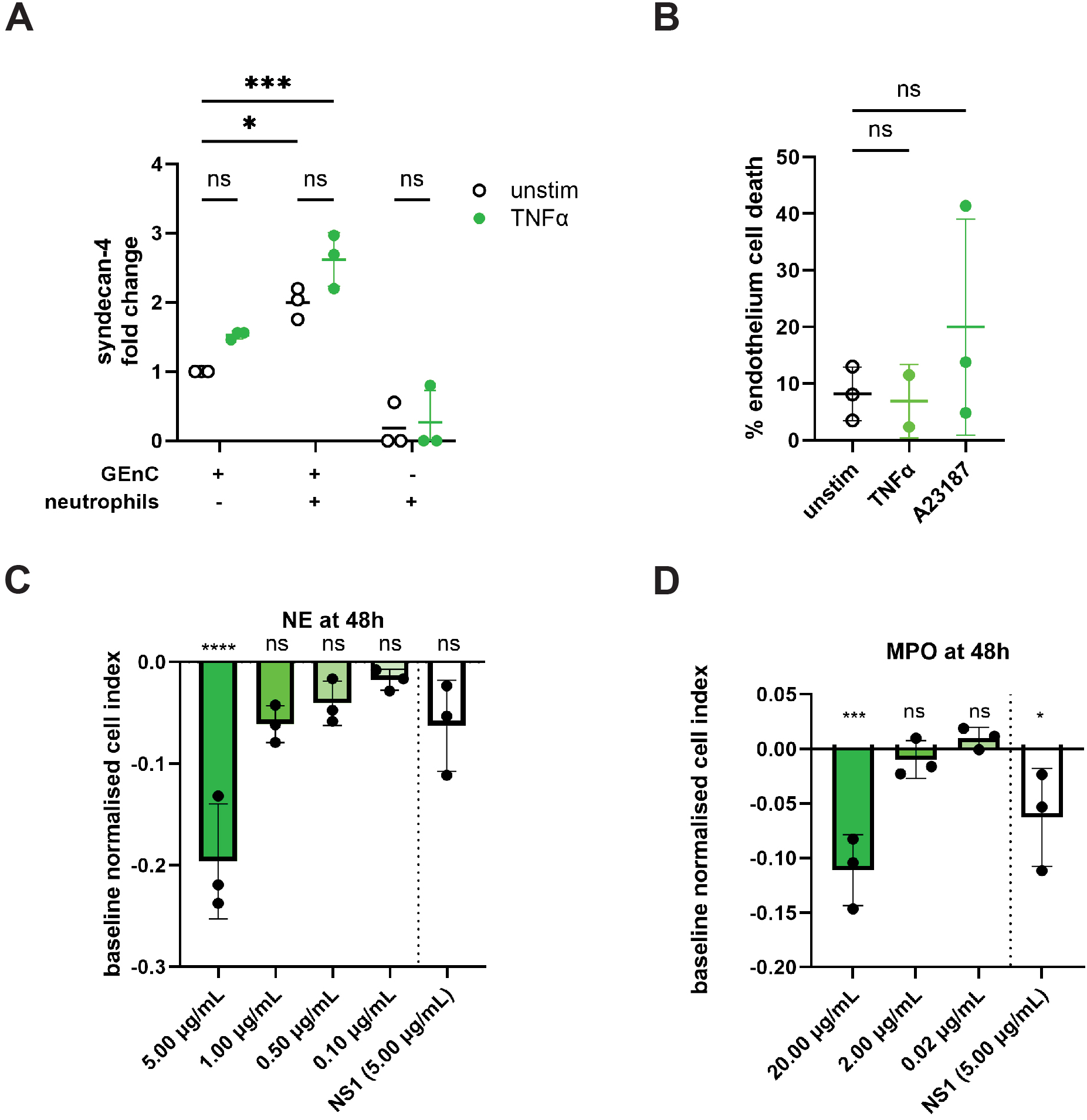
Additional analyses of endothelial permeability and glycocalyx shedding. **A.** Fold change of syndecan-4 in GEnC culture supernatants following co-culture with peripheral blood neutrophils for 5 h, with or without treatment with TNFα (2 ng/mL), n = 3. **B.** Percentage of GEnC death following culture with untreated, TNFα (2 ng/mL) and A23187 (2.5 μM)-treated neutrophils for 5 h, n = 3. **C.** TEER cell index at 48 h following treatment of HUVECs with NE (0.10−5.00 μg/mL) and positive control (Dengue virus NS1 protein; 5.00 μg/mL) and compared to baseline, n = 3. **D.** TEER cell index at 48 h following treatment of HUVECs with MPO (0.02−2.00 μg/mL) and positive control (Dengue virus NS1 protein; 5.00 μg/mL) and compared to baseline, n = 3. Data information: Data are presented as mean ± SD. ns – not significant, \**P* ≤ 0.05, \*\*\**P* ≤ 0.001, \*\*\*\**P* ≤ 0.0001, assessed using two-way ANOVA with Tukey’s post-hoc test (**A**) and one-way ANOVA with Dunnett’s post-hoc test (**B-D**).

**Figure S2.**
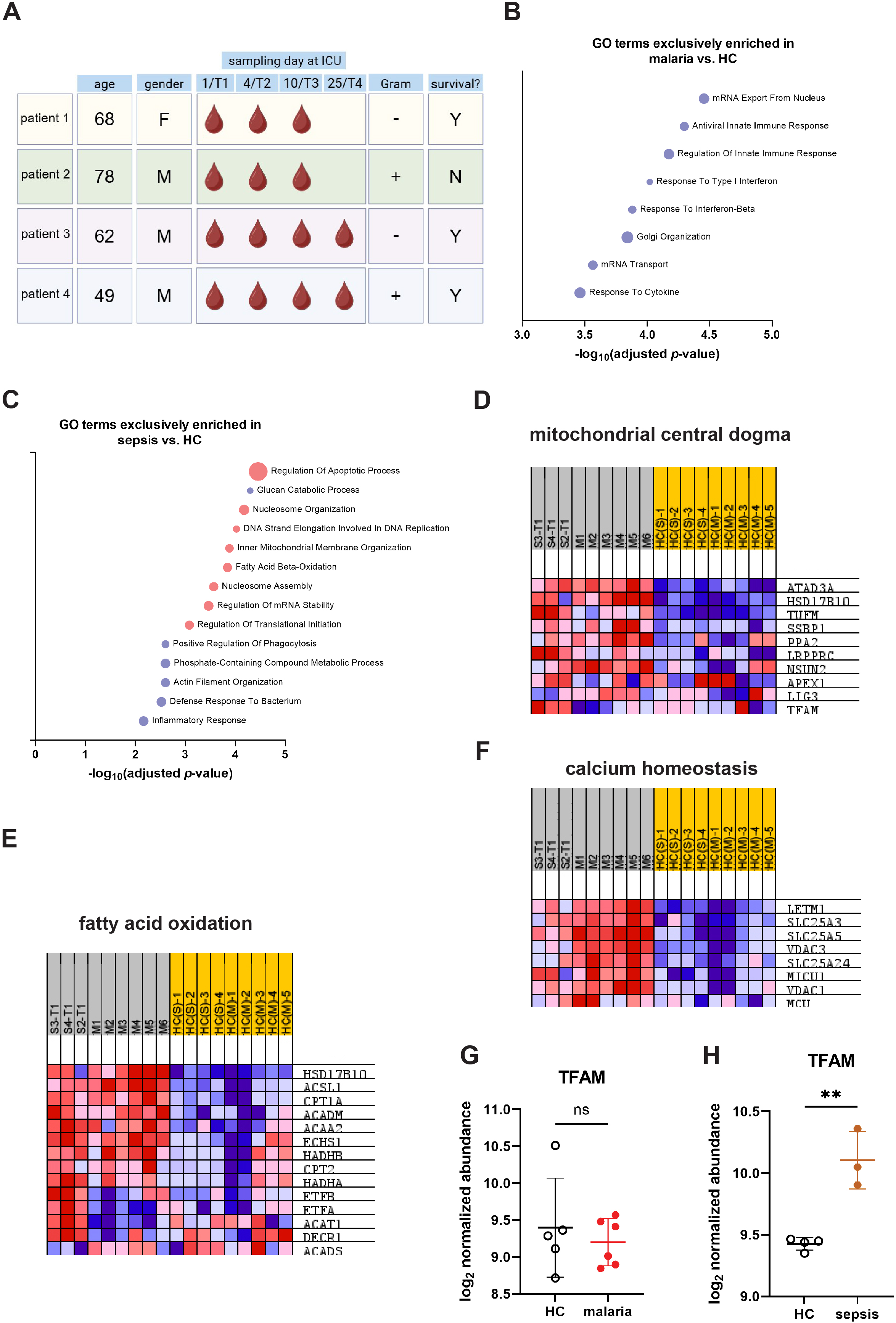
Additional bioinformatics analysis. **A**. Detailed schematic of blood collection from sepsis patients. **B**-**C**. Gene Set Enrichment Analysis (GSEA) results showing GO terms exclusively enriched in malaria vs. HC (B) and sepsis vs. HC (C) datasets. **D**-**F**. GSEA results showing enriched MitoCarta terms. **G**-**H.** Individual normalized abundances of TFAM peptide in malaria vs. HC (G) and sepsis vs. HC (H) neutrophils. Samples from malaria patients were collected at admission; these patients are designated M1–M6. Samples from sepsis patients were collected on days 1, 4, 10 and 25 following admission to the intensive care unit and are designated T1–T4 for each patient (patients S1–S4). Healthy controls on **D**-**F** are indicated HC(S) for sepsis dataset and HC (M) for malaria dataset. Data information: Data are presented as mean ± SD. ns – not significant, \*\**P* ≤ 0.01, assessed using unpaired *t* test (**G, H**).

**Figure S3.**
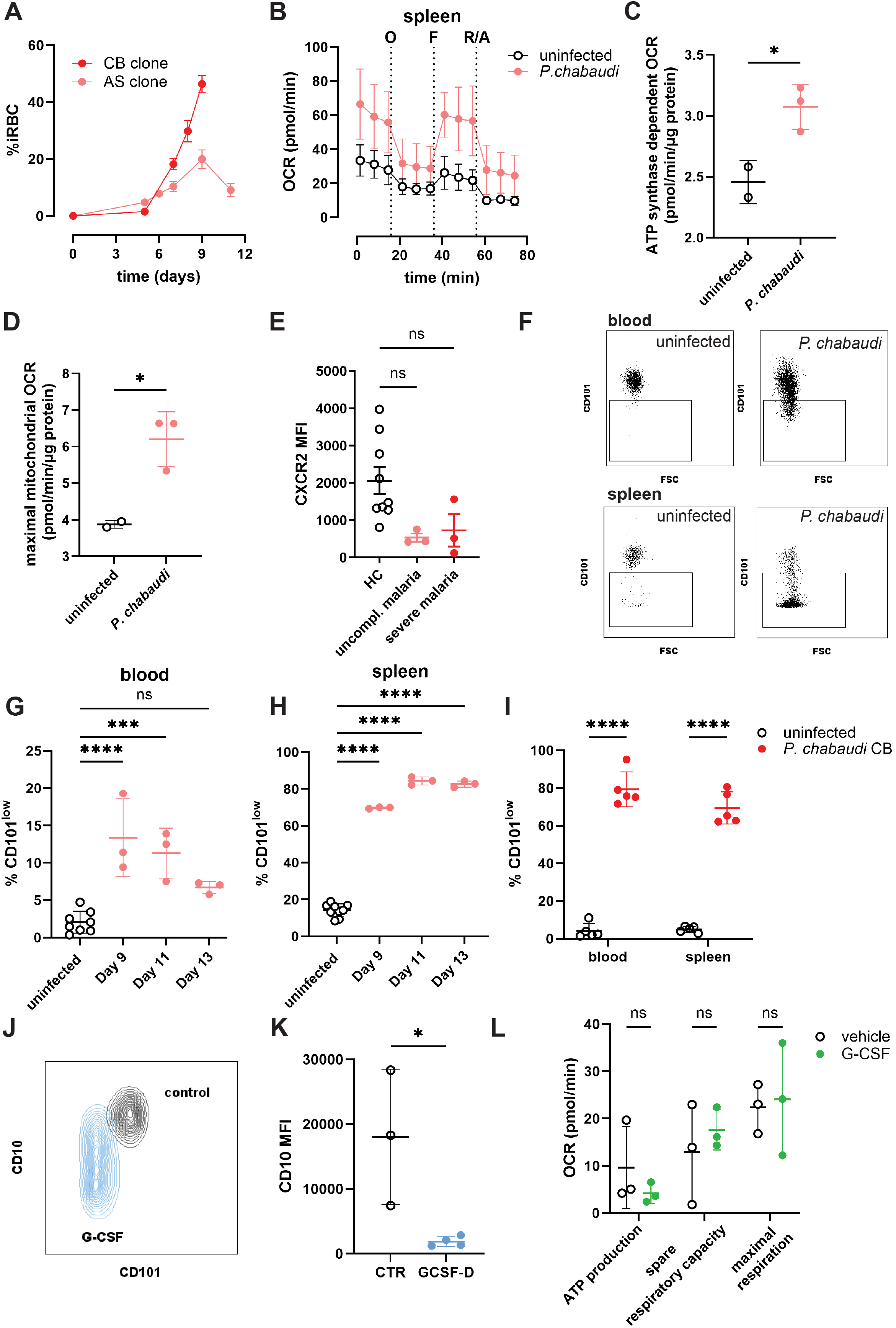
Additional analyses of mitochondrial function in malaria-elicited and GCSF-D neutrophils. **A.** Percentage of infected red blood cells with *P. chabaudi* clone AS (n = 3) and CB (n = 5) in the circulation of mice. **B.** OCR of spleen neutrophils of *P. chabaudi* clone AS-infected mice (n = 3). Seahorse port injections are indicated as: O – oligomycin (1.5 μM), F – FCCP (400 nM), R/A – rotenone (1 μM) and antimycin A (1 μM). **C.** ATP synthase dependent OCR of splenic neutrophils isolated from *P. chabaudi* clone AS-infected mice on day 9 post-infection (n = 3). **D.** Maximal mitochondrial OCR of splenic neutrophils isolated from *P. chabaudi* clone AS-infected mice on day 9 post-infection (n = 3). **E**. Median fluorescence intensity of CXCR2 on peripheral blood neutrophils from HC, and patients with uncomplicated and severe malaria, n = 9 (HC), 3 (uncomplicated malaria), 3 (severe malaria). **F.** Representative flow cytometry plots showing CD101 expression on peripheral blood and splenic neutrophil from mice at day 9 post-infection with *P. chabaudi* clone AS. **G.** Percentage of CD101^low^ peripheral blood neutrophils from *P. chabaudi* clone AS-infected mice at days 9, 11 and 13 post-infection. n = 8 (uninfected mice), 3 (infected mice). **H.** Percentage of CD101^low^ splenic neutrophils from *P. chabaudi* clone AS-infected mice at days 9, 11 and 13 post-infection. n = 8 (uninfected mice), 3 (infected mice). **I.** Percentage of CD101^low^ neutrophils in peripheral blood and spleen from mice infected with *P. chabaudi* clone CB at day 9 post-infection, n = 5. **J.** Representative flow cytometry contour plot for CD10 and CD101 expression on peripheral blood neutrophils derived from control (CTR) and GCSF-treated (GCSF-D) donors. **K.** Median fluorescence intensity of CD10 on peripheral blood neutrophils from CTR and GCSF-D donors, n = 3 (CTR), 4 (GCSF-D). **L.** ATP production, spare respiratory capacity, and maximal respiration of CTR neutrophils treated *ex vivo* with 10 ng/mL G-CSF for 30 min, n = 4. Data information: Data are presented as mean ± SD. ns – not significant, \**P* ≤ 0.05, \*\*\**P* ≤ 0.001, \*\*\*\**P* ≤ 0.0001, assessed using unpaired *t* test (**C**, **D**, **K**), one-way ANOVA with Dunnett’s post-hoc test (**E**, **G, H**), and two-way ANOVA with Šidák’s post-hoc test (**I**, **L**).

**Figure S4.**
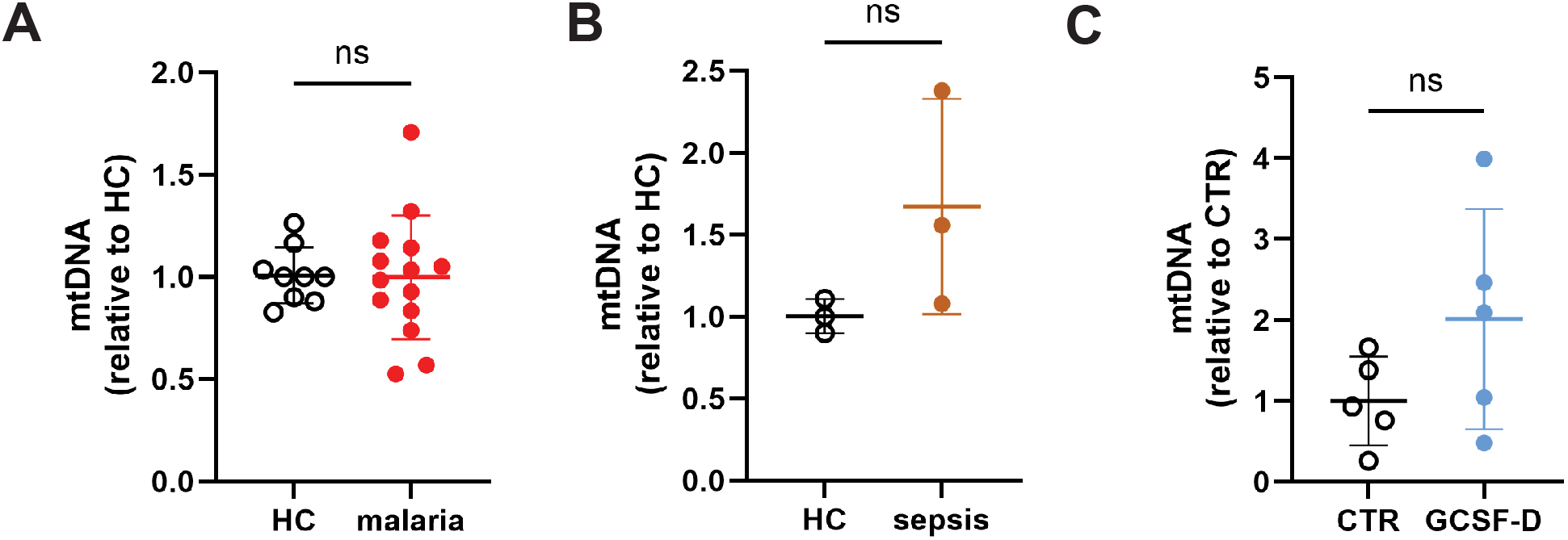
Additional analyses of mitochondria in neutrophils. **A.** Relative mtDNA content in neutrophils from malaria patients (n = 14) compared to HC (n = 9). **B.** Relative mtDNA content in neutrophils from sepsis patients (n = 3) compared to HC (n = 3). **A.** Relative mtDNA content in neutrophils from GCSF-D donors compared to CTR neutrophils, n = 5 each. Data information: Data are presented as mean ± SD. ns – not significant, assessed using unpaired *t* test.

**Fig. S5.**
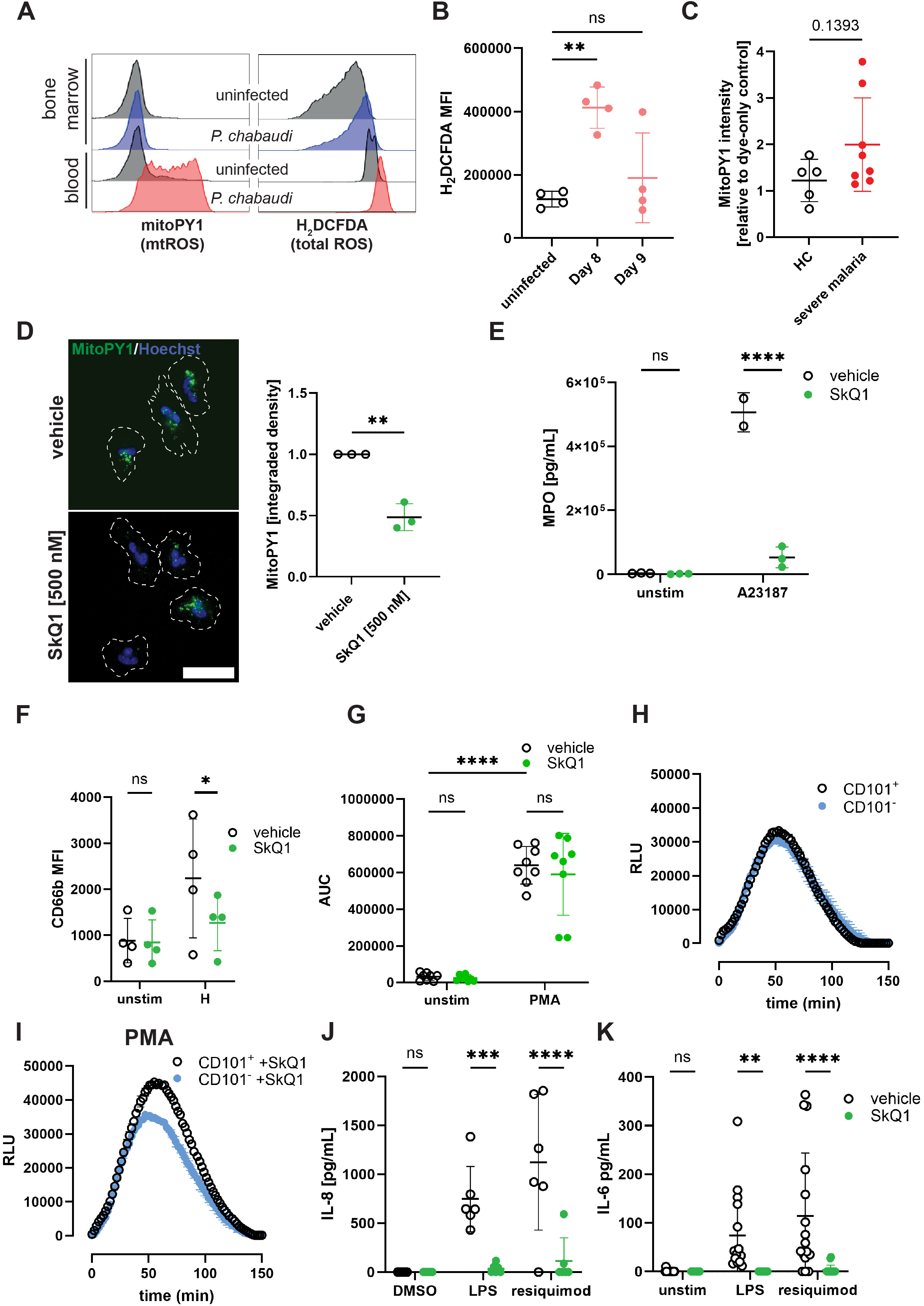
mtROS regulates neutrophil degranulation, NOX2-dependent oxidative burst and cytokine release. **A**. Representative FACS histograms showing mtROS (MitoPY1) and total ROS (H2DCFDA) profiles of blood and bone marrow neutrophils from uninfected and *P. chabaudi* clone AS-infected mouse. **B**. Median fluorescence intensity of H_2_DCFDA in neutrophils from uninfected and *P. chabaudi* clone AS-infected mice at day 8 and 9 post-infection, n = 4. **C**. Fluorescence intensity of MitoPY1 in neutrophils from HC (n = 5) and cerebral malaria patients (n = 8) relative to dye-only control **D**. *Left*: Representative widefield fluorescence images of CTR neutrophils stained with MitoPY1 and Hoechst, pre-incubated with vehicle (DMSO) or SkQ1 (500 nM), scale bar 10 μm. *Right*: MitoPY1 integrated density, n = 3. **E**. Myeloperoxidase (MPO) levels in supernatants of CTR neutrophils stimulated with calcium ionophore A23187 (2.5 μM), pre-incubated with vehicle (DMSO) and SkQ1 (500 nM), n = 3. **F**. Median fluorescence intensity of CD66b on peripheral blood neutrophils from CTR donors stimulated with heme (30 μM), n = 4. **G**. Quantification of oxidative burst (as area under curve, AUC) of CTR neutrophils stimulated with PMA (100 nM), pre-incubated with vehicle (DMSO) and SkQ1 (500 nM) n = 8. **H**. Representative luminol-based measurement of ROS production in FACS-sorted mouse CD101^−^ and CD101^+^ neutrophils stimulated with PMA (100 nM). **I.** Representative luminol-based measurement of ROS production in FACS-sorted mouse CD101^−^ and CD101^+^ neutrophils stimulated with PMA (100 nM) and pre-stimulated with vehicle or SkQ1 (500 nM). **J.** Interleukin 8 (IL-8) levels in supernatants of CTR neutrophils stimulated overnight with LPS (100 ng/mL; n = 6) and resiquimod (5 μM); n = 5), **K**. Interleukin 6 (IL-6) levels in supernatants of CTR neutrophils stimulated overnight with LPS (100 ng/mL) and resiquimod 5 μM), n = 13 (LPS), 15 (resiquimod). Data information: Data are presented as mean ± SD. ns – not significant, \**P* ≤ 0.05, \*\**P* ≤ 0.01, \*\*\**P* ≤ 0.001, \*\*\*\**P* ≤ 0.0001, assessed using one-way ANOVA with Dunnett’s post-hoc test (**B**), unpaired *t* test (**C, D**) and two-way ANOVA with Tukey’s (**E-G**, **J-K**).

**Fig. S6.**
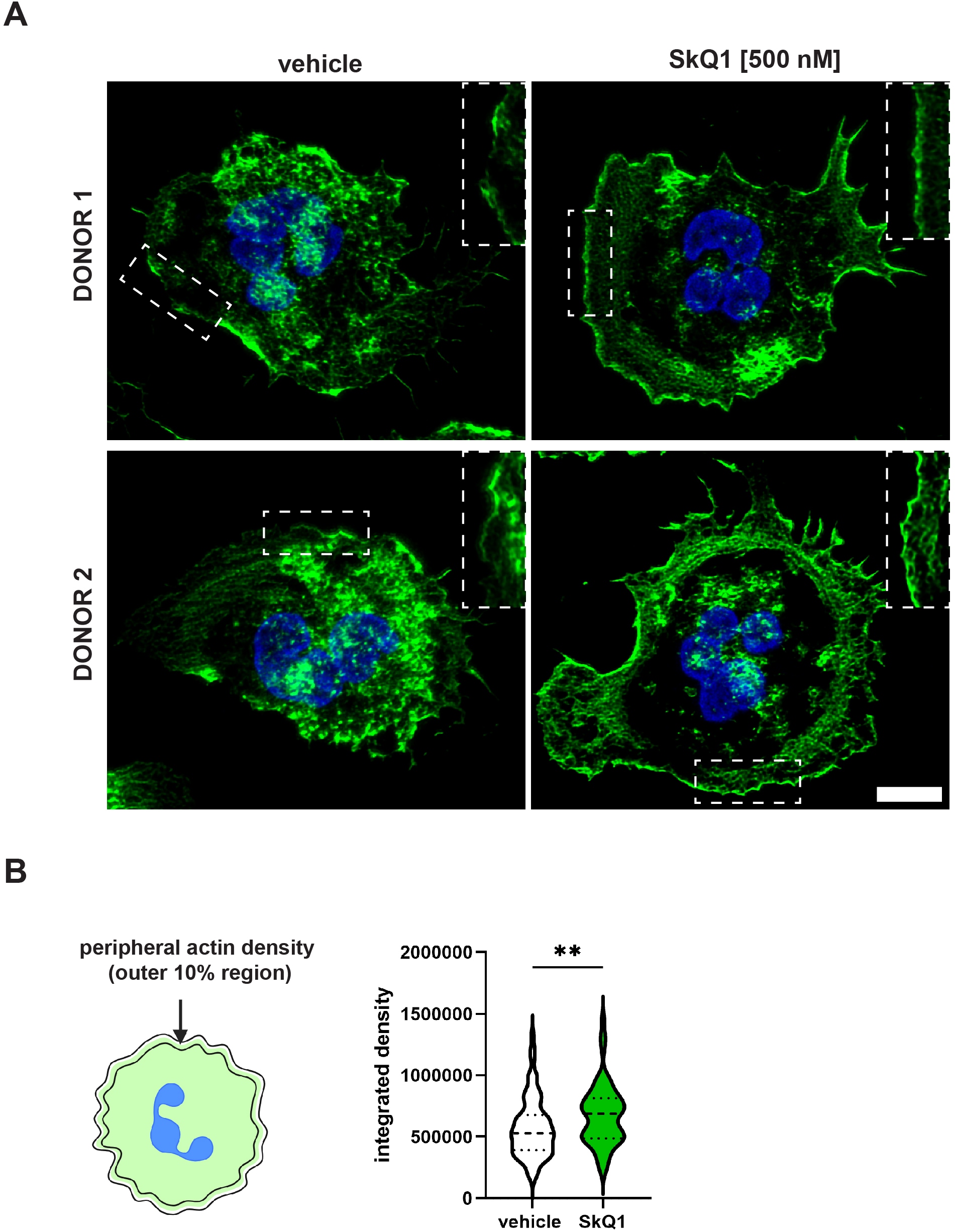
mtROS quenching enhances F-actin accumulation at the cell periphery. **A**. Representative high-resolution confocal images of control neutrophils stained with phalloidin−AlexaFluor^TM^ 488 and DAPI after treatment with vehicle (DMSO) or SkQ1 (500 nM); scale bar 5 μm. **B**. Quantification of phalloidin integrated density in the outer 10% of cell area. Data are shown as pooled single-cell measurements, n = 2 donors/independent experiments. Data information: Data are presented as mean ± SD. \**P* ≤ 0.05, \*\**P* ≤ 0.01, assessed using unpaired Mann-Whitney test (**B**).

